# A VAE-based methodology for deep enterotyping and Parkinson’s disease diagnosis

**DOI:** 10.64898/2026.03.17.26348604

**Authors:** Yuting Qiao, Zhanshan (Sam) Ma

## Abstract

Gut microbiome studies in Parkinson’s disease (PD) are challenged by high dimensionality, sparsity, compositionality, and substantial between-cohort heterogeneity, all of which complicate robust community typing and disease-status classification. Here, we developed a variational autoencoder (VAE)-based methodology for deep enterotyping and PD diagnosis prediction (*i.e*., predicting diseased vs. control status) using a harmonized multi-cohort gut microbiome compendium comprising 1,957 16S rRNA samples from six PD case–control cohorts and an independent shotgun metagenomic validation cohort of 725 samples. Compared with conventional enterotyping approaches such as partitioning around medoids (PAM) and Dirichlet multinomial mixture (DMM) modelling, the VAE-derived latent space supported a clearer and more reproducible three-cluster solution. These three enterotype-like community states were biologically interpretable and were annotated as Enterococcus-type, Bacteroides-type, and Ruminococcus-type configurations. The same broad three-enterotype structure was independently recapitulated in the metagenomic dataset, supporting cross-platform robustness. Across the three inferred types, the proportion of PD samples was similar, and both the primary generalized linear mixed-effects model and sensitivity model showed that enterotype assignment was not a significant differentiating factor for PD status and that the lack of association was not dependent on a single modelling strategy. In the supervised branch, VAE-derived representations supported PD case–control classification while also providing a shared latent representation for clustering, enterotype transfer, and downstream interpretation. Collectively, these findings show that deep representation learning can improve the resolution, reproducibility, and interpretability of enterotype inference in heterogeneous microbiome datasets, and provide a practical methodology for organizing broad community structure in PD. In this setting, the main advantage of the VAE method lies in its ability to link unsupervised community typing with supervised prediction through a shared latent representation, even when broad community types do not function as stand-alone disease biomarkers.

## 1. Introduction

The human gut microbiome is a densely populated and highly complex ecological system that varies markedly across individuals and populations. This variation reflects the combined influence of host physiology, environmental exposures, diet, lifestyle, and technical factors associated with sampling and sequencing, resulting in microbiome datasets that are high-dimensional, sparse, and compositional. Consequently, a recurring goal in microbiome research is to derive community-level representations that are both interpretable and robust, enabling meaningful comparisons across cohorts and supporting downstream analyses that link microbiome structure to host phenotypes and disease outcomes.

To address this challenge, the concept of enterotypes was proposed as recurrent gut community configurations that stratify individuals into a limited number of compositional clusters (Arumugam et al., 2011). Enterotypes were originally framed as broad community patterns dominated by specific taxa and accompanied by functional differences, offering a practical abstraction for organizing inter-individual heterogeneity. Subsequent work has extended enterotyping toward translational contexts, including the use of enterotype-like groupings as candidate biomarkers and as a basis for stratifying individuals with respect to clinical outcomes and intervention responsiveness (Costea et al., 2018). Recent examples further suggest that enterotypes may capture clinically relevant microbial configurations—for instance, a Megamonas-dominated enterotype has been robustly associated with obesity in a large-scale cohort, underscoring the potential utility of community typing as a disease-relevant descriptor (Rao et al., 2024). A similar principle has been observed in other host-associated microbiomes: in the vaginal microbiome, community states may reflect both phenotype and community complexity rather than a single disease-aligned canonical type, and machine-learning models can nevertheless classify these states with high precision, underscoring the translational potential of microbiome community typing while also highlighting its dependence on how community structure is defined (Ma, 2023).

In neurodegenerative disorders such as Parkinson’s disease (PD), there is substantial interest in identifying reproducible microbiome patterns that could reflect biologically meaningful host–microbe interactions, support patient stratification, and help clarify heterogeneity in disease manifestation. PD is clinically diverse, and gastrointestinal features, diet, medication, and lifestyle differences may all shape gut community structure, making confounding and cohort heterogeneity particularly salient. This concern is supported by recent large-scale reanalyses showing that PD-associated gut microbiome alterations are strongly scale-dependent across cohorts: alpha-diversity differences appear only in a subset of comparisons, whereas broader beta- and gamma-diversity patterns remain more informative at the community level (Qiao & Ma, 2026). These findings reinforce the view that PD microbiome structure is heterogeneous and may not be adequately summarized by single taxa or by simple diversity indices alone. In this setting, enterotype-style community typing is attractive because it can provide a compact representation of gut community organization that is potentially more stable than individual taxa. However, robust inference is essential: community types that primarily reflect confounders (e.g., diet or bowel habits) rather than disease-related signals would hinder mechanistic interpretation and translational value. This concern is consistent with the broader enterotype literature emphasizing that host and environmental factors can strongly influence enterotype structure and its stability (Vandeputte et al., 2015; Wu et al., 2011).

Despite their conceptual appeal, enterotypes remain controversial, in part because the existence and number of discrete clusters can be method- and cohort-dependent. Several studies have argued that gut microbiome variation is often better represented as continuous gradients (“enterogradients”) rather than sharply separated classes, and that cluster boundaries can be weak or unstable across datasets (Knights et al., 2014; Costea et al., 2018). Consistent with this view, recent ecological analyses in PD suggest that dysbiosis may be expressed more through altered stochasticity, composition, and species interaction structure than through a wholesale reorganization of canonical community categories, implying that disease-related signal may be distributed and only partially recoverable by classical discrete clustering alone (Qiao & Ma, 2025). Moreover, critical evaluations highlight that seemingly canonical structures (e.g., a three-enterotype scheme) are not consistently reproduced, and that the inferred number of clusters can vary with preprocessing decisions, distance metrics, and clustering algorithms (Cheng & Ning, 2019). These observations imply that enterotyping is not only a biological question but also a methodological one: whether “types” emerge depends strongly on how one represents and partitions compositional community profiles.

From a computational standpoint, enterotype inference is an unsupervised clustering problem performed on sparse, compositional abundance data. Classical approaches include partitioning around medoids (PAM), typically using Jensen–Shannon divergence, which is intuitive and robust to outliers but requires the user to predefine the number of clusters *K* and may be sensitive to the choice of distance metric (Arumugam et al., 2011). Dirichlet multinomial mixture (DMM) models provide a probabilistic alternative that can select an “optimal” cluster number via model selection criteria (e.g., BIC) and handle sparsity, but they may overfit in small samples and rely on distributional assumptions (Holmes et al., 2012). Beyond hard clustering, topic-model formulations such as latent Dirichlet allocation (LDA) represent each microbiome as a mixture of latent assemblages, offering higher-resolution structure and partial memberships, though with reduced ease of assigning discrete labels and potential instability across initializations (Hosoda et al., 2020). More recent covariate-aware methods, exemplified by ENIGMA, incorporate host variables into the generative model to disentangle latent community structure from phenotype-associated shifts, directly addressing enterotype–phenotype confounding at the modeling level (Abe et al., 2019).

However, the limitations of existing pipelines are particularly consequential when the goal is to test enterotype–disease relationships in heterogeneous cohorts such as PD. First, many clustering-based methods provide cluster assignments without principled uncertainty estimates and are vulnerable to sensitivity in *K* selection: if *K* is too small, distinct patterns may be conflated; if too large, redundant or spurious subtypes may be produced. Second, confounders can be “absorbed” into the clustering structure: apparent enterotypes may reflect systematic differences in diet, sampling, medication, or other covariates rather than latent ecological states. While ENIGMA demonstrates how enterotype-like clustering can be integrated into association analysis, its Bayesian hierarchical structure introduces additional model complexity and hyperparameters, and may face scalability and reproducibility issues when data are sparse or when priors/initializations lead to different local optima (Abe et al., 2019).

A further and increasingly recognized concern is over-optimism in unsupervised microbiome analysis. Because unsupervised workflows often involve many reasonable analytical choices (normalization, filtering, distances, algorithms, and parameter settings), exploring multiple pipelines and selectively reporting the most “compelling” result can inflate perceived robustness and hinder reproducibility. Systematic examinations of this phenomenon indicate that clustering outcomes can appear stable in discovery data yet fail to replicate in validation datasets, especially when the analysis implicitly optimizes toward a desired structure (Ullmann et al., 2023). These issues motivate best practices emphasizing transparent sensitivity analyses, stability checks, and automated or consensus-based state-definition pipelines that reduce the risk of spurious clusters (García-Jiménez & Wilkinson, 2019). For enterotype research aimed at disease association, these safeguards are not optional: they are necessary to prevent the inadvertent conversion of cohort structure or confounding into “biological” community types.

In parallel, advances in representation learning suggest an opportunity to revisit enterotyping with deep generative models. Classical clustering typically operates on relative abundances or transformed profiles, but such representations can be noisy and sensitive to sparsity and compositional effects. By contrast, deep latent-variable models can learn low-dimensional embeddings that denoise community profiles and capture nonlinear structure that may be difficult to represent with conventional distances. Related work in computational biology has used neural-network-based approaches to identify archetypal spaces or latent structures in complex datasets, illustrating the broader potential of deep learning for data-driven subtyping (van Dijk et al., 2019).

Moreover, deep-learning-based clustering has begun to show value for endotyping and response classification in multi-omic contexts, highlighting the feasibility of integrating deep representation learning with clustering to derive clinically meaningful subgroups (Rockel et al., 2025). More broadly, AI- and machine-learning-based microbiome analyses across major neurodegenerative, neurodevelopmental, and psychiatric disorders have already demonstrated strong diagnostic feasibility, but these efforts have largely focused on supervised species-based discrimination rather than robust unsupervised representation of community states (Ma et al., 2025). These developments motivate the hypothesis that deep embeddings may enable more reproducible microbiome community typing—provided that model selection, stability, and confounding are handled rigorously.

Here, we present a variational autoencoder (VAE)-based deep enterotyping methodology coupled with *k*-means clustering, designed to improve robustness and interpretability in microbiome community typing and to enable accurate prediction of Parkinson’s disease (PD) diagnosis—by which we mean computational classification of diseased versus control samples based on gut microbiome profiles, not clinical diagnosis. Our approach targets two persistent challenges in the enterotype literature: robust selection of the number of community types (*K*) and confounder adjustment when relating community types to disease status. Specifically, the VAE learns a compact latent representation of microbiome profiles (from 16S rRNA gene sequencing and, where available, metagenomic feature representations such as functional gene cluster profiles), after which k-means partitions the latent space into candidate community types. We incorporate confounder correction within the modeling and/or association-testing method to reduce the risk that inferred enterotypes merely reflect non-disease covariates, aligning with the motivation of covariate-aware enterotyping models such as ENIGMA (Abe et al., 2019). Finally, to address concerns about over-optimism and instability in unsupervised analysis, we evaluate performance and robustness using five-fold cross-validation and explicit stability criteria, and we benchmark our method against established classical enterotyping pipelines, including PAM and DMM (Arumugam et al., 2011; Holmes et al., 2012).

By integrating deep representation learning with robust clustering and confounder-aware inference, our methodology aims to provide a reproducible enterotyping approach that is well-suited for PD microbiome studies. We further use the inferred community types to test whether enterotype-like structure is associated with PD status after adjusting for relevant covariates, thereby clarifying the extent to which community typing can capture disease-related microbiome variation beyond major confounding gradients. Collectively, this work seeks to strengthen the methodological foundations of enterotyping and to enable more reliable investigation of microbiome–disease relationships in Parkinson’s disease.

## 2. Materials and Methods

### 2.1. Datasets and generation of taxonomic abundance tables

We assembled a multi-cohort gut microbiome compendium for Parkinson’s disease (PD) comprising seven independent case–control datasets, including six 16S rRNA amplicon cohorts and one shotgun metagenomic cohort. Dataset-level metadata (BioProject accession, sample size, and country/year of collection) were curated from the original publications. The 16S compendium included 1,957 samples (PD = 1,153, controls = 804) from six BioProjects: PRJDB8639 (Japan, 2020), PRJEB27564 (Finland, 2019), PRJEB30615 (Germany, 2019), PRJNA381395 (Germany, 2019), PRJNA601994 (USA, 2020), and PRJNA742875 (Korea, 2021). The metagenomic cohort comprised 725 samples (PD = 490, controls = 235) from PRJNA834801 (USA, 2019). The metagenomic dataset was used exclusively for external validation; specifically, we derived genus-level abundance profiles from shotgun reads using a consistent processing pipeline and used these profiles as an independent validation input, whereas the primary benchmarks were conducted on the harmonized 16S genus-level matrix.

For 16S rRNA amplicon data, raw paired-end reads were retrieved from NCBI and processed using a unified taxonomic profiling pipeline to derive five-rank abundance tables (phylum, class, order, family, and genus). Sequencing adapters and low-quality bases were removed using Trimmomatic (v0.39) with standard filtering parameters, and the resulting high-quality reads were subjected to taxonomic classification using Kraken2 (v2.1.2) with the SILVA reference database. Read-level classifications were subsequently re-estimated into abundance profiles using Bracken (v2.6) to improve rank-specific quantification. The per-sample Kraken reports were converted into consistent tabular formats with KrakenTools, and rank-specific feature matrices were assembled in R (v4.1.2) by merging all samples on a unified taxonomy index. Each rank-specific table was generated by (*i*) extracting the corresponding rank from Bracken outputs, (*ii*) sorting taxonomy identifiers to enforce consistent row ordering across samples, and (*iii*) concatenating sample-wise abundance vectors to form an abundance matrix (Fig 1). (Bolger et al., 2014; Wood et al., 2019; Lu et al., 2017; Quast et al., 2013).

**Figure 1.**
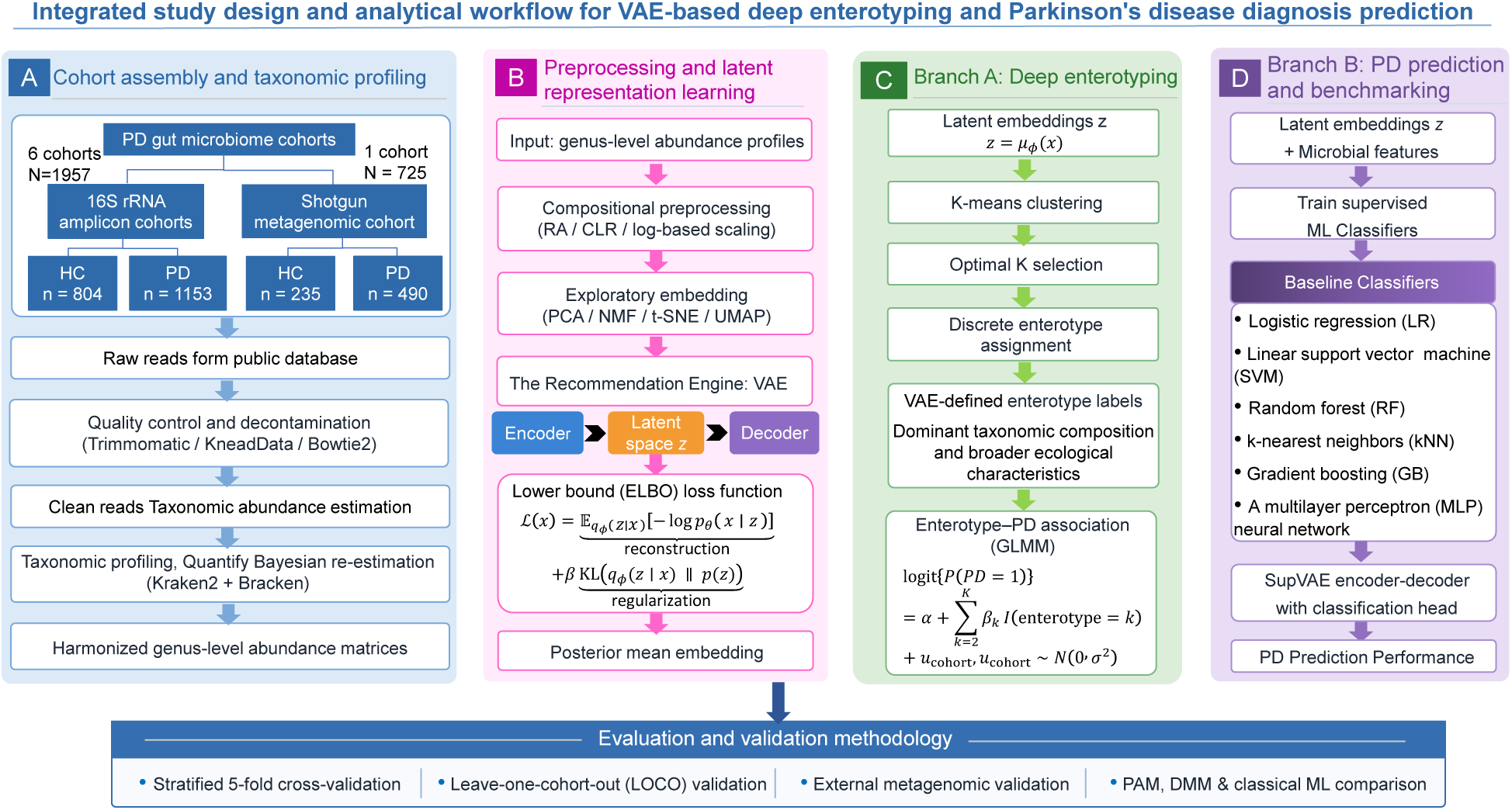
Integrated study design and analytical workflow for VAE-based deep enterotyping and Parkinson’s disease diagnosis prediction: Public PD gut microbiome cohorts, including six 16S rRNA amplicon datasets and one shotgun metagenomic dataset for external validation, were processed through a unified taxonomic profiling pipeline to generate harmonized genus-level abundance matrices. After compositional preprocessing and transformation screening, microbiome profiles were embedded into a VAE-derived latent space. The latent representation was then used in two downstream branches: (A) deep enterotyping, where k-means clustering and silhouette-guided model selection were used to define discrete enterotypes and assess their association with PD by GLMM; and (B) PD diagnosis, where supervised latent representations and selected microbial features were used for classification and benchmarked against conventional machine-learning models. Model robustness and generalizability were evaluated using stratified five-fold cross-validation, leave-one-cohort-out validation, and external metagenomic validation, with additional comparison against classical enterotyping methods including PAM and DMM.

For shotgun metagenomic profiling, preprocessing and host decontamination were performed using the EasyMetagenome workflow (v1.11) in a Linux environment. Briefly, reads were quality controlled and de-hosted using KneadData with Trimmomatic trimming and Bowtie2 alignment against a pre-built Homo sapiens reference index, retaining only non-host reads for downstream analyses (Bolger et al., 2014; Langmead & Salzberg, 2012). Taxonomic classification was performed with Kraken2 against a local Kraken2 PlusPF database, and abundance re-estimation was conducted with Bracken using the corresponding Bracken database derived from the same reference (Fig 1) (Wood et al., 2019; Lu et al., 2017). To ensure comparability with the 16S benchmarks, we constructed the same five-rank abundance tables (phylum, class, order, family, genus) by aggregating Bracken estimates at each rank and merging per-sample outputs with a consistent taxonomy ordering. As quality control, host-related lineages were removed when detected (e.g., excluding Chordata/Metazoa and residual human assignments). The resulting metagenome-derived genus-level table retained bacterial as well as archaeal, fungal, and viral genera when classified by the reference database, enabling multi-kingdom genus-level profiling.

In this study, these genus-level profiles were used as an independent external validation input for evaluating cross-dataset generalizability of the proposed models.

### 2.2. VAE latent representation learning for enterotyping and PD diagnosis

We implemented a two-branch method that shares a variational autoencoder (VAE) encoder for representation learning. The learned latent vector 𝑧 is then used for (*i*) deep enterotyping via k-means clustering and (*ii*) PD classification via supervised predictors trained on latent embeddings.

This design yields a unified latent space that supports both unsupervised community typing and disease diagnosis while enforcing a consistent evaluation protocol (Fig 1).

#### 2.2.1. Preprocessing of compositional microbiome profiles

Microbiome abundance profiles are inherently sparse and compositional. To reduce the impact of library-size differences and to account for compositional constraints, we considered five commonly used transformations prior to training the variational autoencoder (VAE) and subsequent community-typing analyses.

First, we evaluated relative abundance (L1 normalization) by converting each sample’s OTU/taxon counts to proportions (each feature divided by the total counts per sample), a standard normalization step in microbial ecology to improve comparability across samples (Lozupone & Knight, 2008). Second, we applied the centered log-ratio (CLR) transformation, which is designed for compositional data: a small pseudocount was added to all entries to avoid undefined log values, and the log-transformed abundances were centered by subtracting the sample-wise geometric mean (equivalently, subtracting the mean of log-transformed values within each sample) (Aitchison, 1982). In addition, we assessed three log-based scaling strategies commonly used in machine-learning workflows: log1p followed by StandardScaler (zero mean, unit variance), log1p followed by RobustScaler (median centering and IQR scaling), and log1p followed by MinMax scaling to the [0, 1] range (Pedregosa et al., 2011).

To screen for a suitable preprocessing option for unsupervised community typing, each transformed table was embedded using four complementary dimensionality-reduction methods: PCA (Ringnér, 2008), NMF (Lee & Seung, 1999), t-SNE (van der Maaten & Hinton, 2008), and UMAP (McInnes et al., 2018). For each preprocessing–embedding combination, we clustered samples using k-means and selected the number of clusters 𝐾 by maximizing the average silhouette coefficient over a predefined candidate range (typically 𝐾 = 2–10; extended up to 15 in sensitivity checks) (Rousseeuw, 1987). In addition to the maximizing criterion, silhouette curves and cluster-size distributions were inspected to avoid non-informative solutions (e.g., highly imbalanced or near-empty clusters). This exploratory procedure was used only to guide the choice of preprocessing; all downstream model evaluations were conducted on held-out folds under the cross-validation protocols described below to limit optimistic bias.

#### 2.2.2. VAE architecture and objective

Let 𝑥 ∈ ℝ* denote the transformed microbiome feature vector. The encoder parameterizes a diagonal Gaussian approximate posterior:

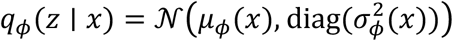

where 𝜇,(𝑥)and log 𝜎^8^(𝑥)are parameterized by a feed-forward neural network. Latent variables are sampled using the reparameterization trick:

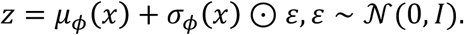

The decoder defines 𝑝_C_(𝑥 ∣ 𝑧)and produces a reconstruction 𝑥. Model training optimizes a 𝛽-regularized evidence lower bound (ELBO):

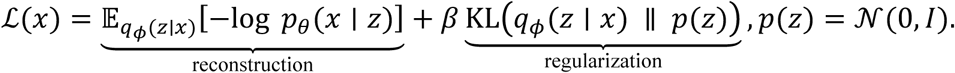

where 𝛽 ≥ 0 controls the strength of latent regularization. These objective balances accurate reconstruction of microbiome profiles with a structured latent space suitable for clustering (Kingma & Welling, 2014; Higgins et al., 2017).

#### 2.2.3. Architecture and training configuration

We implemented a fully connected VAE with a 2-dimensional latent space. The encoder comprised two dense layers and produced the latent mean and log-variance vectors 𝜇*_ϕ_*(𝑥) and 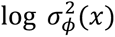. The posterior mean 𝜇*_ϕ_*(𝑥) was used as a deterministic embedding for downstream clustering.

For downstream analyses, we used the posterior mean 𝜇*_ϕ_*(𝑥) as a deterministic embedding and denote it by 𝑧 ≡ 𝜇*_ϕ_*(𝑥) rather than using stochastic samples from 𝑞*_ϕ_*(𝑧 ∣ 𝑥) . This choice improves reproducibility of embeddings across resampling and cross-cohort validation and enables consistent nearest-centroid assignment in held-out data.

### 2.3. Branch A: VAE-based deep enterotyping

In this study, we adopted a discrete enterotype method, treating gut community types as separable states and assigning each sample to a single enterotype label (hard assignment). After representation learning with the VAE encoder, each sample was mapped to a latent vector 𝑧. Enterotype labels were derived by k-means clustering in latent space, and test samples were assigned by nearest-centroid mapping to ensure consistent hard assignments under cross-validation.

k-means clustering was performed on the latent embeddings with candidate cluster numbers evaluated over a predefined range (𝐾 = 2–20). For each candidate 𝐾, clustering was fitted on the training embeddings, and the resulting centroids were used to assign samples (including held-out samples) by nearest-centroid mapping. This hard-assignment formulation provides discrete community labels that can be used for downstream robustness assessment and association testing. The optimal number of enterotypes (𝐾^∗^ ) was selected by maximizing the average silhouette coefficient, with 𝐾^∗^ determined within the training split only to avoid information leakage. To reduce spurious solutions driven by outliers or degenerate partitions, we additionally inspected cluster-size distributions and applied a cautionary minimum cluster-size threshold (5% of samples) during interpretation.

To assess the stability and generalizability of discrete enterotypes, we implemented a two-layer evaluation strategy: (*i*) Unsupervised stability. We performed stratified five-fold cross-validation (preserving PD/control proportions) and leave-one-cohort-out (LOCO) validation. Within each training split, preprocessing, VAE training, 𝐾-selection, and k-means fitting were performed exclusively on the training data. Held-out samples were embedded using the fixed encoder and assigned to the nearest training-set centroid. Because cluster labels are permutation-invariant, labels were aligned across folds using the Hungarian algorithm prior to computing agreement. Stability was quantified using cross-fold clustering agreement (ARI/NMI), held-out silhouette scores, and cluster-size dispersion. In addition, bootstrap resampling was used to compute membership-level Jaccard stability. (*ii*) Supervised predictability (auxiliary validation). To evaluate whether inferred discrete enterotypes correspond to learnable and reproducible structure in latent space (rather than unstable partitions), we trained an auxiliary multilayer perceptron classifier on latent embeddings to predict enterotype labels under the same five-fold and LOCO protocols. Permutation testing (1,000 permutations) was used to estimate empirical 𝑝-values for the observed predictive performance. This auxiliary task was used as a consistency check of latent-space separability.

### 2.4. Enterotype–PD association analysis with confounder control

To test whether inferred enterotype-like community types were associated with Parkinson’s disease (PD) while accounting for between-study heterogeneity, we fitted a generalized linear mixed-effects model (GLMM) with a logit link, modelling PD status as a function of enterotype membership and including cohort (BioProject) as a random intercept:

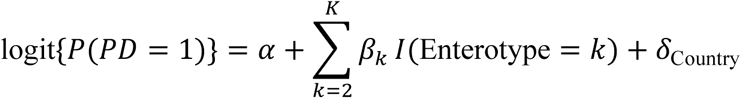

Here, 𝐼(enterotype = 𝑘) denotes indicator variables for enterotype membership, with one enterotype specified as the reference category. Odds ratios (ORs) and 95% confidence intervals (CIs) were obtained by exponentiating model coefficients. Because country of recruitment was highly correlated with cohort structure in this dataset, country was evaluated in a sensitivity analysis using a fixed-effects logistic regression model rather than included in the primary mixed-effects model.

Because country of recruitment was highly correlated with cohort structure in this dataset, country was not included in the primary mixed-effects model to avoid overadjustment and collinearity. Instead, country was evaluated in a sensitivity analysis using a fixed-effects logistic regression model:

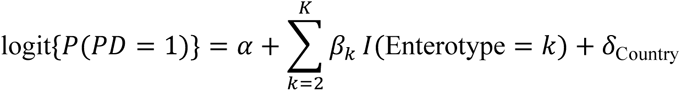

where 𝛿_Country_ represents country-specific fixed effects. This sensitivity analysis was used to assess whether the inferred association between enterotype and PD was robust to adjustment for broad geographic background.

The overall contribution of enterotype was assessed using likelihood ratio tests comparing models with and without the enterotype term.

### 2.5. Branch B: PD diagnosis using SupVAE representations and machine learning classifiers

To evaluate the predictive utility of the learned microbiome representation, we formulated Parkinson’s disease (PD) diagnosis as a binary classification task (PD vs. healthy control). Starting from the transformed microbiome abundance matrix described above, we implemented a supervised variational autoencoder (SupVAE) method in which latent representation learning and disease discrimination were optimized jointly. For each sample, the encoder produced a deterministic latent representation defined by the posterior mean, 𝑧 = 𝜇,(𝑥), which was then used for downstream classification.

Because preliminary analyses showed that latent variables alone were insufficient to maximize discrimination, the final diagnosis branch used a hybrid representation that combined the latent embedding 𝑧with a subset of microbiome features selected from the transformed input matrix. Within each training split, features were first filtered by prevalence and total abundance, after which the transformed matrix was subjected to univariate feature ranking and the top 𝐾 features were retained. These selected features were concatenated with the latent representation to form the final classifier input. In the optimized model, 𝐾 = 200.

The SupVAE consisted of a fully connected encoder that parameterized the latent mean and log-variance, and a mirrored decoder used for reconstruction. A supervised classification head was attached to the latent mean and jointly optimized with the reconstruction and KL terms. The training objective was defined as the sum of reconstruction loss, KL divergence, and classification loss:

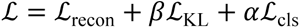

where 𝛽 controls latent regularization and 𝛼 controls the contribution of the classification term. During model tuning, we evaluated multiple latent dimensions, 𝛽 values, 𝛼 values, selected-feature numbers, and downstream classifiers.

For downstream diagnosis, we compared several candidate classifiers during model development, including logistic regression, gradient boosting, multilayer perceptron, and random forest. Based on cross-validation performance, Random Forest was retained as the final classifier for the SupVAE-based hybrid representation.

We also evaluated a variant that incorporated enterotype information into the diagnosis branch. To avoid information leakage, enterotype labels for held-out samples were not taken directly from the metadata during cross-cohort validation. Instead, within each training split, latent centroids were computed for the enterotype groups using the training data only and held-out samples were assigned hard enterotype labels by nearest-centroid mapping in the training latent space. These reassigned enterotype labels were one-hot encoded and concatenated with 𝑧and the selected microbiome features before Random Forest classification.

Model development and parameter selection were first carried out using stratified five-fold cross-validation, preserving PD/control proportions in each fold. Within each fold, all preprocessing steps, feature filtering, feature selection, SupVAE fitting, and downstream classifier training were performed exclusively on the training split, and the held-out fold was used only for evaluation. Model performance was summarized using ROC-AUC as the primary metric, together with accuracy, sensitivity, specificity, balanced accuracy, and F1 score.

To assess cross-cohort generalizability, we additionally performed strict leave-one-cohort-out (LOCO) validation. In each iteration, one cohort was held out as the test set and all remaining cohorts were used for training. All preprocessing, feature filtering, feature selection, SupVAE training, latent embedding, enterotype reassignment, and classifier fitting were restricted to the training cohorts. Decision thresholds for binary classification were not fixed a priori; instead, they were selected within the training data by an inner train/validation split to maximize balanced accuracy and then applied unchanged to the held-out cohort. This procedure ensured that both probability estimation and threshold calibration were determined without access to the test cohort.

### 2.6. Benchmark enterotyping and diagnosis with classical methods

#### 2.6.1. Partitioning Around Medoids (PAM)

We applied PAM (k-medoids) clustering to genus-level profiles. The optimal number of clusters 𝑘was determined by maximizing the Calinski–Harabasz (CH) index over candidate solutions 𝑘 = 1–20. Cluster compactness and separation were quantified using the average silhouette coefficient. To visualize between-sample dissimilarities and cluster structure, ordination was performed using principal coordinates analysis (PCoA) based on Jensen–Shannon divergence (JSD), followed by between-class analysis (BCA) to emphasize separation among inferred enterotypes.

#### 2.6.2. Dirichlet Multinomial Mixture (DMM)

We fitted Dirichlet multinomial mixture (DMM) models to the same genus-level abundance table to obtain a probabilistic, model-based community typing. Model selection was conducted by scanning candidate component counts (typically 𝑘 = 1 –20) and choosing the solution that minimized the Laplace-approximated model fit criterion, yielding the final number of Dirichlet components. To assess the global arrangement and overlap among DMM-derived community types, we visualized samples using non-metric multidimensional scaling (NMDS) based on Bray–Curtis dissimilarity, with samples colored by inferred enterotype labels. Dominant genera driving each component was summarized via a feature heatmap. To probe phenotype-linked heterogeneity, DMM models were additionally fitted separately to healthy controls and PD cases, and the optimal component count was recorded for each group.

#### 2.6.3. Baseline classifiers

We benchmarked a panel of standard machine-learning classifiers commonly used for microbiome-based disease diagnosis, including logistic regression (LR), linear support vector machine (SVM), random forest (RF), k-nearest neighbors (kNN), gradient boosting (GB), and a multilayer perceptron (MLP) neural network (NN). For each cohort, microbial abundance tables from controls and cases were read as floating-point feature matrices and concatenated into a single sample-by-feature matrix. Binary class labels were assigned as 0 for controls and 1 for cases. Prior to model training, features were z-score standardized using StandardScaler to obtain zero-mean, unit-variance transformed abundances.

## 3. Results

### 3.1. Conventional enterotype models showed limited structure in the merged PD microbiome dataset

#### 3.1.1. Partitioning around medoids (PAM) revealed weak cluster separation

We first applied partitioning around medoids (PAM) to the merged dataset of 1,957 gut microbiome profiles to evaluate whether conventional distance-based clustering could recover discrete enterotype structure. The optimal number of clusters was selected using the Calinski–Harabasz (CH) index across candidate values of 𝐾, and the highest CH value was observed at 𝐾 = 3, indicating that a three-cluster solution was most strongly supported under this criterion (Fig. 2A).

Based on the conventional naming strategy in enterotype analysis, in which clusters are labeled according to the genus showing the highest relative abundance within each cluster, the three PAM-derived clusters were designated as Bacteroides, Ruminococcus–Faecalibacterium, and Ruminococcus–Blautia enterotypes. Visualization of the cluster assignments in principal component analysis (PCA) space showed that the three groups could be distinguished at a broad level, with the Bacteroides cluster occupying a relatively separate region and the two Ruminococcus-related clusters distributed on the opposite side of the ordination (Fig. 2B). A between-class analysis (BCA) projection further highlighted the three-cluster configuration, but also showed that samples remained connected by gradual transitions rather than forming sharply bounded groups (Fig. 2C).

**Figure 2A.**
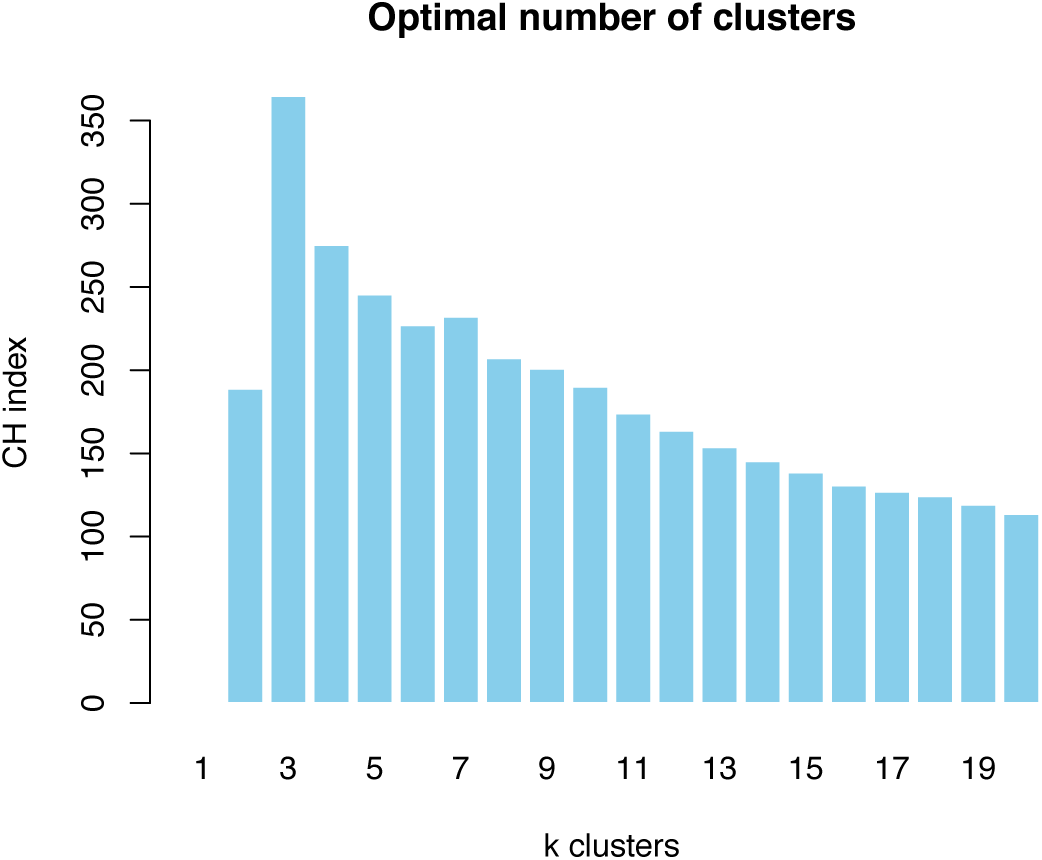
Selection of the optimal number of PAM clusters using the Calinski–Harabasz index. The Calinski–Harabasz (CH) index was calculated across candidate cluster numbers for partitioning around medoids (PAM). The highest CH value was observed at 𝐾 = 3, supporting a three-cluster solution for downstream enterotype analysis.

**Figure 2B.**
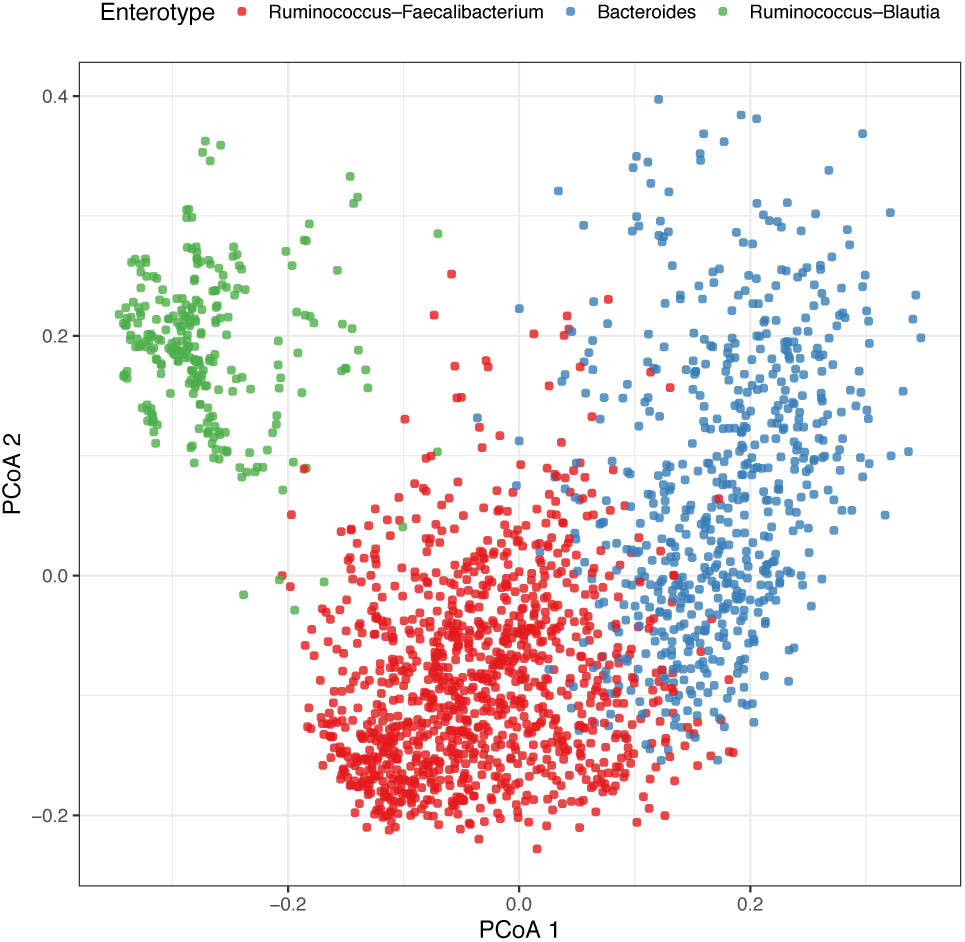
PAM-derived enterotype assignments visualized in principal component analysis space. Samples were clustered by PAM and projected onto the first two principal components. Colors indicate the three enterotypes identified by PAM, labeled according to the dominant genus or co-dominant enrichment pattern within each cluster: Ruminococcus–Faecalibacterium, Bacteroides, and Ruminococcus–Blautia.

**Figure 2C.**
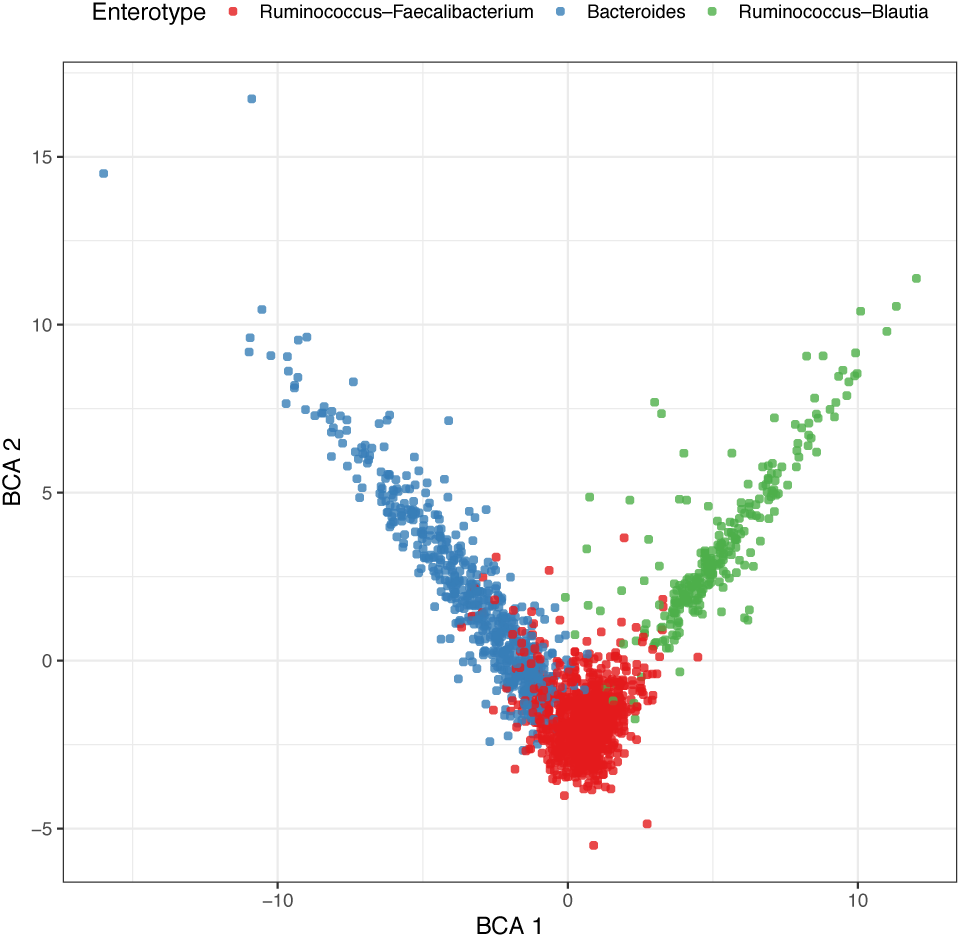
Between-class analysis of PAM-derived enterotypes. The three PAM-derived enterotypes were visualized using between-class analysis (BCA). Samples are colored according to enterotype assignment. The projection shows broad separation among the three clusters, while also indicating continuous transitions and partial overlap between groups.

The overall separation of the PAM-derived clusters was limited. The mean silhouette coefficient for the three-cluster solution was 0.149, indicating weak compactness and substantial overlap among clusters despite the apparent three-group structure. Thus, although PAM recovered a nominal three-enterotype partition, the resulting boundaries were diffuse and the cluster structure remained only moderately resolved.

#### 3.1.2. DMM showed a continuous enterotype structure

Dirichlet multinomial mixture (DMM) modelling supported a 12-component solution for the merged dataset, as the Laplace criterion reached its minimum at *k* = 12 among the tested models (Fig. 3A). This result indicates that, relative to simpler partitioning approaches, the gut microbiome profiles in the combined dataset can be decomposed into a larger number of latent community components.

Pairwise examination of DMM component weights showed that correlations among the inferred components were uniformly low and predominantly negative, generally ranging from approximately −0.04 to −0.15 (Fig. 3B). Such a pattern suggests that these components were not fully independent or sharply separated states, but rather partially exclusive mixture structures distributed within a shared compositional landscape.

**Figure 3A.**
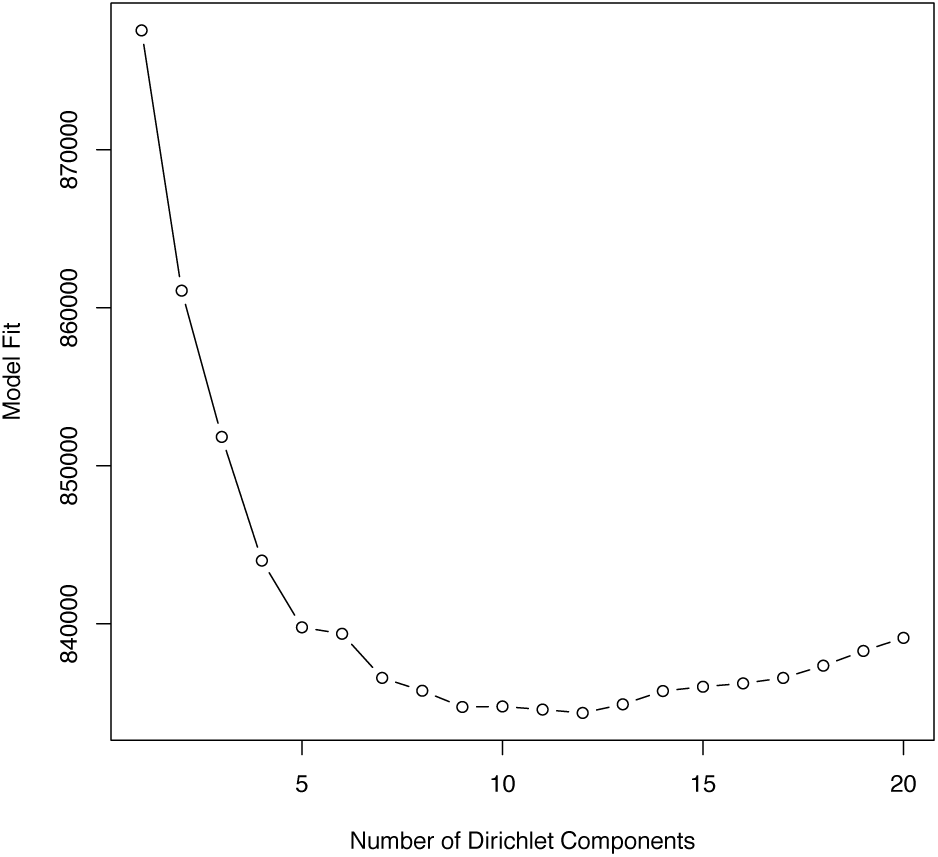
Model selection using the Laplace criterion identified a 12-component DMM solution as the optimal fit.

**Figure 3B.**
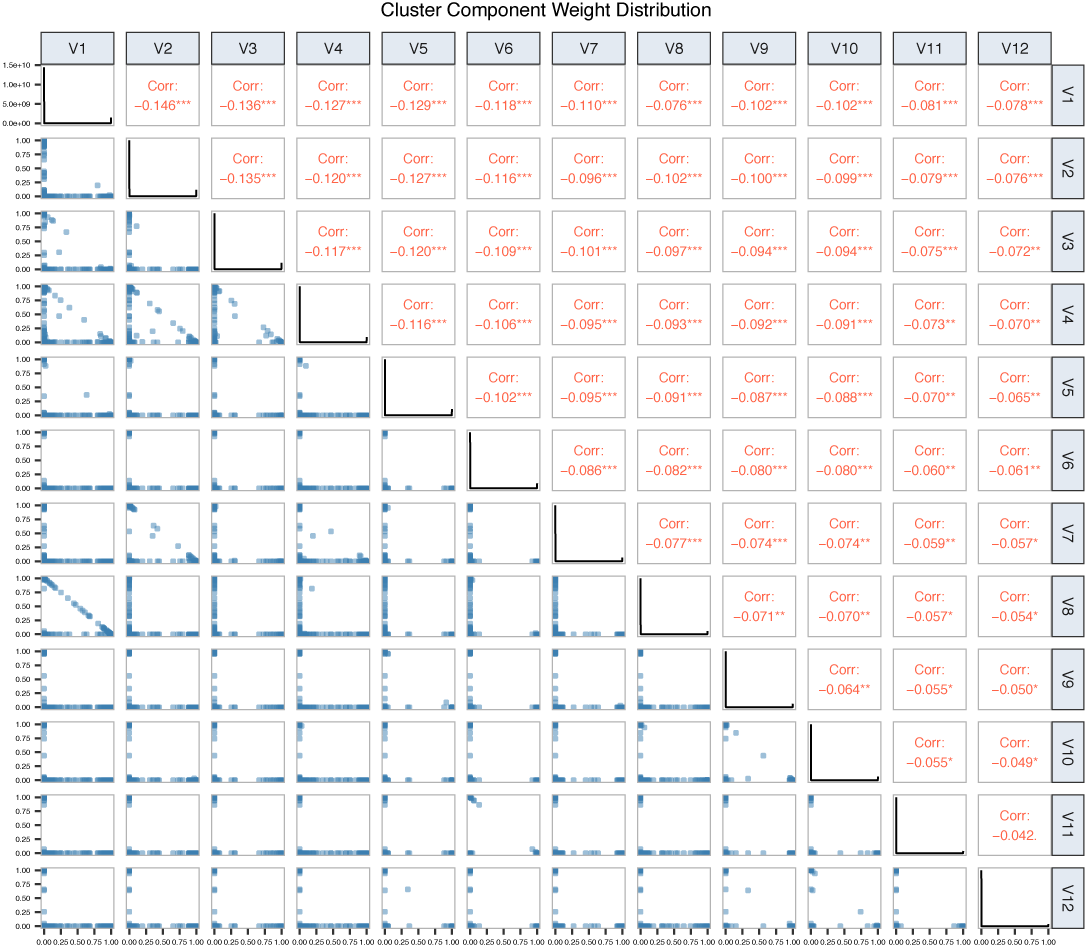
Pairwise distributions and correlation coefficients of DMM component weights showed uniformly weak negative correlations among components, indicating partial exclusivity with substantial overlap in compositional structure.

This interpretation was further supported by the ordination pattern. In NMDS space, the 12 DMM-derived enterotypes showed only partial separation and substantial overlap, despite some local aggregation of samples belonging to the same component (Fig. 3C). Thus, although DMM produced a more refined subdivision than PAM, the inferred enterotype structure still appeared to follow a continuous compositional gradient rather than a set of clearly discrete microbiome states.

**Figure 3C.**
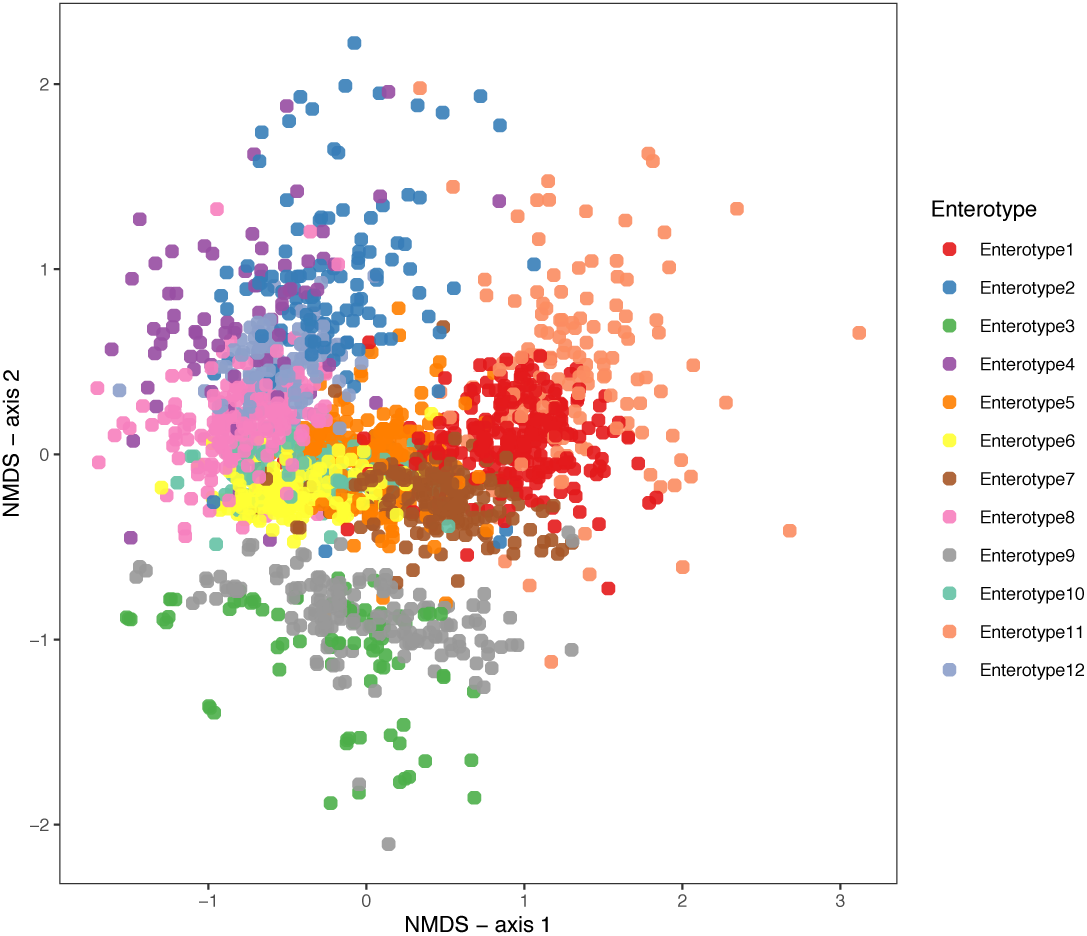
NMDS ordination of samples coloured by DMM-defined enterotype showed partial clustering but extensive overlap among the 12 inferred groups, consistent with a largely continuous community gradient.

The genus-level heatmap yielded a consistent picture (Fig. 3D). Across the 12 DMM-defined enterotypes, Bacteroides, Ruminococcus, and Faecalibacterium remained the principal contributors in most groups, whereas differences among enterotypes were mainly reflected by variations in the relative contributions of secondary genera, including Clostridium, Blautia, Oscillospira, Klebsiella, Lachnospira, Parabacteroides, Akkermansia, Bifidobacterium, Lactobacillus, Coprococcus, Prevotella, Dorea, Collinsella, Eubacterium, and Streptococcus. These results indicate that the DMM solution captured finer-scale heterogeneity within a broadly shared taxonomic background, rather than defining twelve fully discrete canonical enterotypes.

**Figure 3D.**
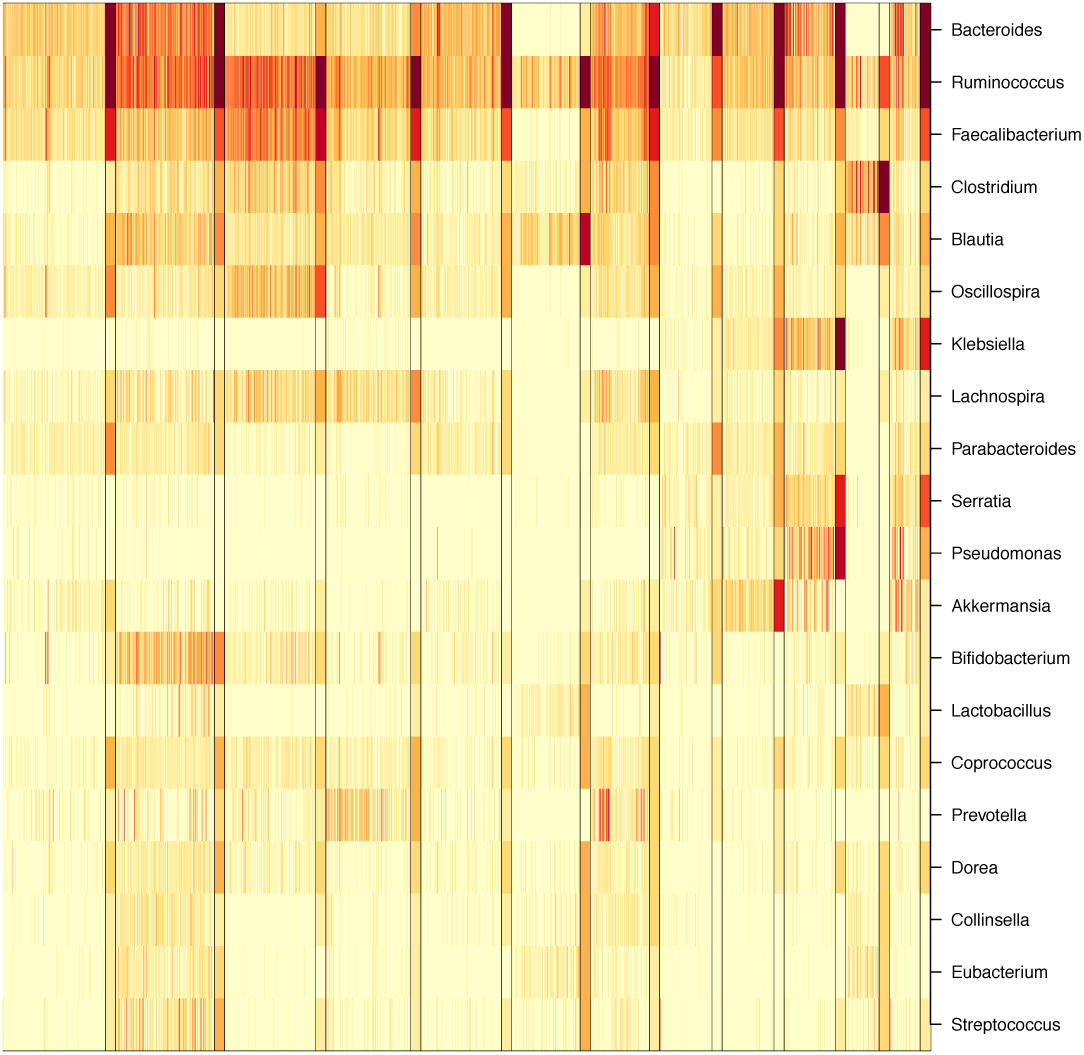
Genus-level heatmap of the 12 DMM enterotypes revealed recurrent dominance of *Bacteroides*, *Ruminococcus*, and *Faecalibacterium*, while differences among enterotypes were mainly driven by shifts in secondary genera.

In addition, phenotype-stratified summary statistics showed that the Healthy and PD groups differed in their optimal component numbers and classification performance (Table 1). The Healthy subset was best fitted by 7 components, whereas the PD subset required 10 components, suggesting greater compositional complexity in PD. The corresponding classification accuracy was 67.1% for Healthy samples and 78.2% for PD samples, with an overall accuracy of approximately 73.5%. Although discrimination was modest, the slightly higher accuracy in PD implies that some DMM-defined community configurations were more frequently represented in PD than in controls. Overall, these findings show that DMM provided a more granular description of microbiome heterogeneity than PAM, but the resulting community structure remained only partially separated and weakly associated with disease status. Table 1 reports the optimal number of DMM components (*k*), sample size, number of taxa, Laplace value, and classification accuracy for the Healthy and PD subsets. The PD subset required more mixture components than the Healthy subset, indicating greater internal compositional heterogeneity.

**Table 1.** Summary of DMM model complexity and classification performance in Healthy and PD groups.

| Group | k | Samples | Taxa | Laplace | Accuracy |
| --- | --- | --- | --- | --- | --- |
| Healthy | 7 | 804 | 510 | 336362.8 | 67.1% |
| PD | 10 | 1153 | 510 | 504641.5 | 78.2% |

### 3.2. VAE-based latent representation improved enterotype separation and supported robust k-means clustering

As an initial benchmark, we projected the L1-normalized relative-abundance profiles using four commonly used dimensionality-reduction methods, including PCA, NMF, t-SNE, and UMAP, and performed k-means clustering with the optimal cluster number determined by silhouette analysis; however, these conventional embeddings produced variable optimal *K* values and only limited or uneven sample separation (Fig. S1). By comparison, the VAE-derived latent representation showed substantially improved clustering structure relative to PAM, DMM, and conventional low-dimensional embeddings, providing clearer enterotype boundaries and more stable support for discrete k-means assignment.

We next developed a variational autoencoder (VAE)-based method to resolve enterotype structure in the merged 16S dataset. Examination of the 1,957-sample amplicon dataset showed that taxon representation followed an approximately log-normal, long-tailed distribution, with a large number of low-abundance features and a smaller set of highly represented taxa (Fig. 4A). This distribution is consistent with the expected sparsity pattern of gut microbiome data and supported the removal of extremely rare taxa before downstream modelling.

After compositional preprocessing by L1 normalization, CLR transformation, and robust scaling, samples were projected into a VAE-derived latent space and clustered using k-means. Silhouette analysis across candidate cluster numbers identified *K* = 3 as the optimal solution, with the highest average silhouette score (0.396) observed at this value (Fig. 4B). This result supported a stable three-cluster partition of the dataset.

**Figure 4A.**
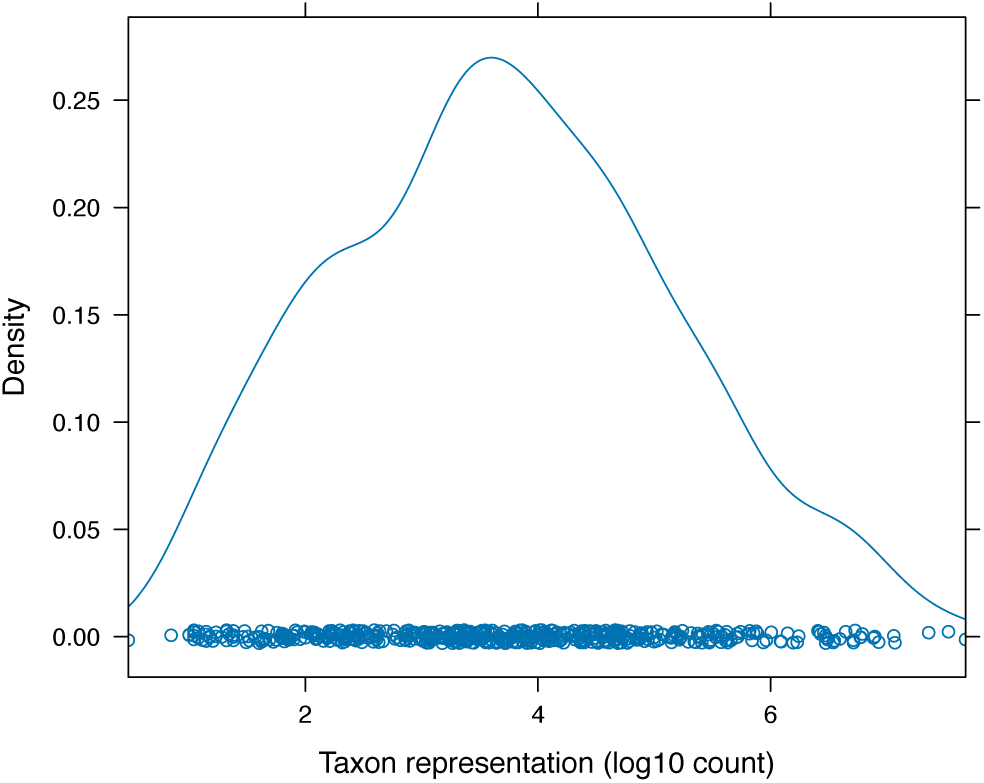
Distribution of taxon representation across the 1,957-sample amplicon dataset, showing an approximately log-normal, long-tailed abundance structure with many low-abundance features.

**Figure 4B.**
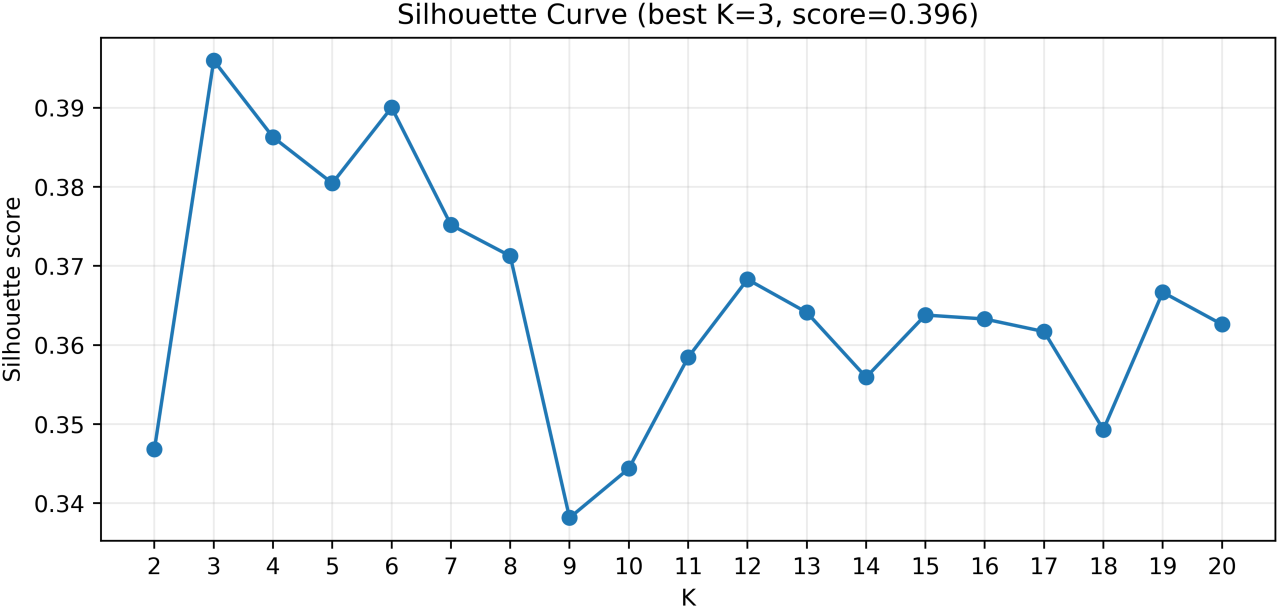
Silhouette analysis for VAE-based enterotyping of 16S rRNA microbiome profiles. Genus-level profiles from the 16S compendium (N = 1,957) were preprocessed using the centered log-ratio (CLR) transformation and embedded into a two-dimensional latent space using a variational autoencoder (VAE). k-means clustering was then applied to the 2D latent embeddings across a range of candidate cluster numbers (𝐾), and clustering quality was evaluated using the average silhouette coefficient. The silhouette scores are shown to support selection of the optimal number of enterotypes.

In the two-dimensional latent space, the three k-means clusters showed visibly improved separation relative to the substantial overlap observed with PAM, DMM, and conventional low-dimensional embeddings (Fig. 4C; Fig. S1). Although some boundary mixing remained, the VAE-derived latent representation produced a markedly clearer clustering structure, indicating that the nonlinear latent space captured microbiome variation more effectively than conventional distance-based or shallow embedding approaches.

**Figure 4C.**
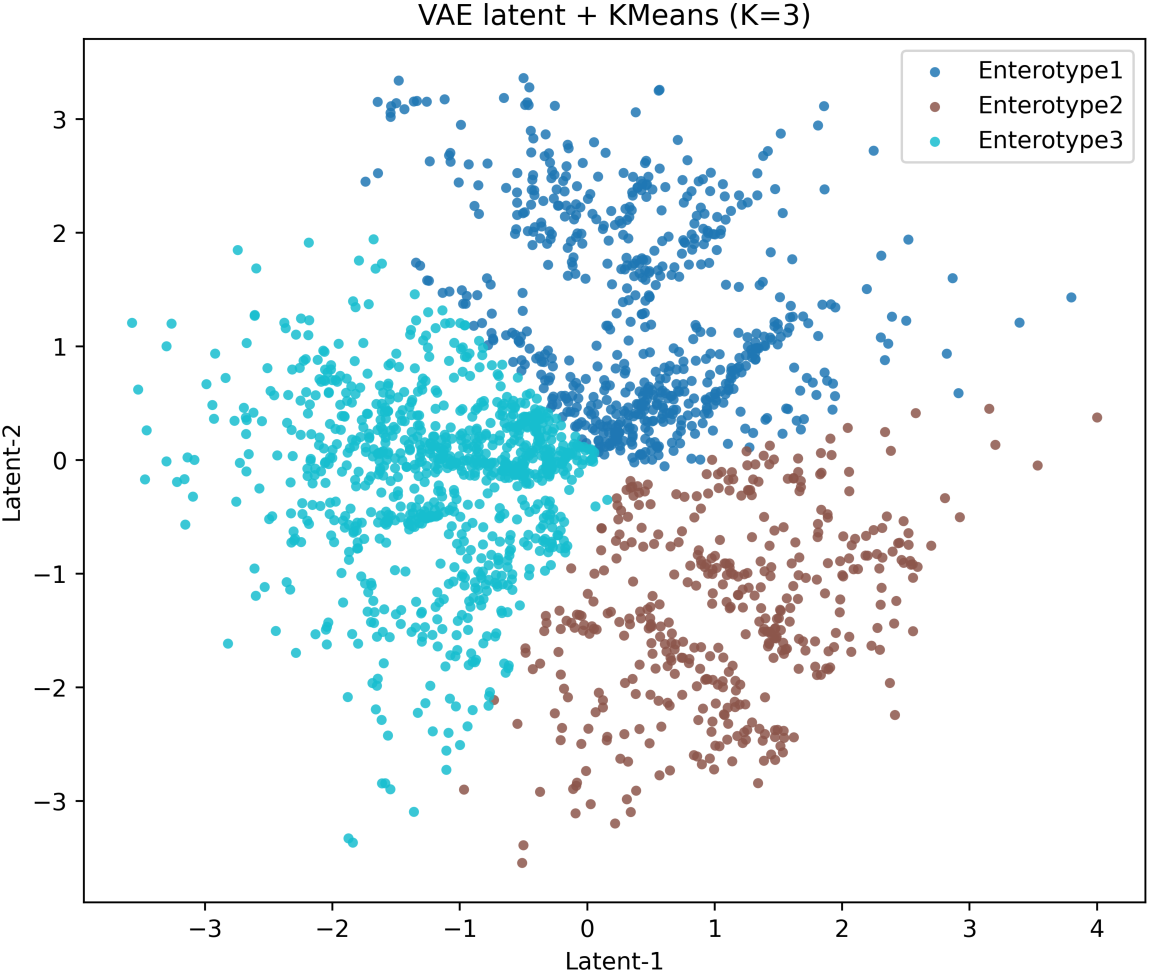
VAE-based enterotyping of 16S rRNA microbiome profiles after CLR transformation. Genus-level profiles from the 16S compendium (N = 1,957) were preprocessed using the centered log-ratio (CLR) transformation and embedded into a two-dimensional latent space with a variational autoencoder (VAE). Samples were then clustered by k-means with 𝐾 = 3, and the resulting cluster assignments are visualized in the 2D latent space to illustrate the inferred enterotype structure.

The three inferred enterotypes contained 626, 417, and 914 samples, respectively (Fig. 4D). Both PD and healthy samples were represented in all three clusters, indicating that the enterotypes reflected broad community configurations rather than disease-specific classes. Nonetheless, the unequal distribution of samples among clusters suggested that some microbiome states were more prevalent than others in the combined dataset.

**Figure 4D.**
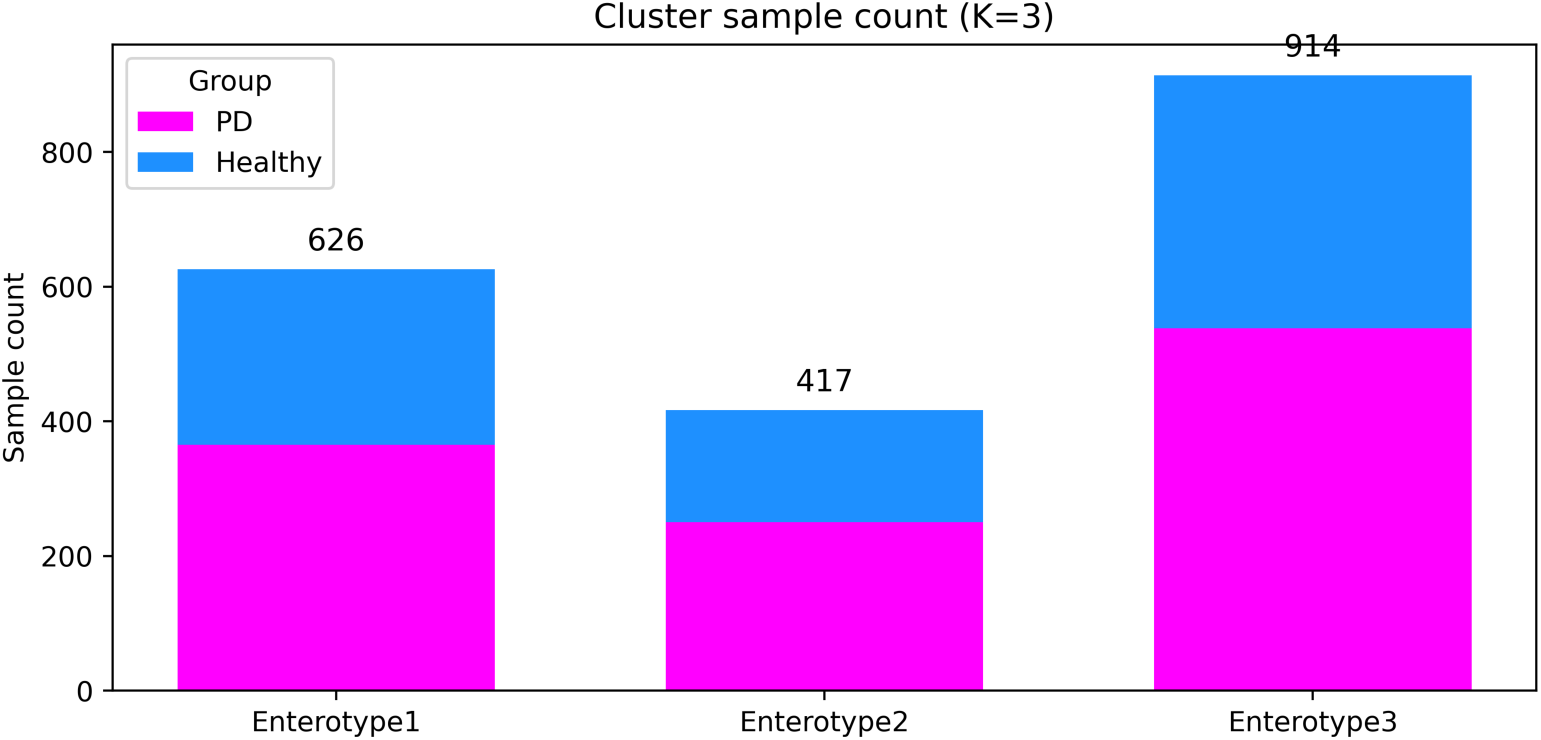
Distribution of case–control sample counts across inferred clusters of 16S rRNA microbiome profiles. A bar chart shows, for each cluster, the number of control (healthy) and PD (disease) samples assigned to that cluster based on the k-means enterotype labels.

Based on their dominant genera and ecological characteristics, the three VAE-defined clusters were annotated as an Enterococcus-type (E-type), Bacteroides-type (B-type), and Ruminococcus-type (R-type). The E-type corresponded to a relatively dysbiosis-like configuration characterized by reduced diversity and enrichment of opportunistic taxa, consistent with a stress- or inflammation-associated community state. The B-type was dominated by *Bacteroides* and represented a bile-tolerant, protein- and fat-associated community configuration. The R-type was characterized by *Ruminococcus* enrichment and was broadly consistent with a fiber-degrading, short-chain fatty acid-producing configuration, although part of this cluster also suggested a more inflammatory substate associated with increased mucin degradation and amino acid fermentation. Taken together, these results indicate that the VAE-based latent representation provided a more biologically interpretable and structurally stable enterotype solution than PAM, DMM, or conventional low-dimensional embeddings.

To assess whether the VAE-based enterotype structure was reproducible across data types, we applied the same workflow to the metagenomic dataset comprising 725 samples. The species-count distribution showed a bimodal and right-skewed pattern, consistent with the coexistence of many low-representation taxa and a smaller set of more abundant taxa, and therefore supported filtering of low-prevalence and low-abundance features before clustering (Fig. S2A).

After L1 normalization, CLR transformation, robust scaling, and abundance/prevalence filtering, silhouette analysis again identified *K* = 3 as the optimal clustering solution, with the highest average silhouette score at 0.354 (Fig. S2B). In the VAE-derived latent space, the three clusters showed discernible, although not complete, separation (Fig. S2C). The three metagenomic enterotypes comprised 209, 377, and 139 samples, respectively (Fig. S2D).

Notably, this three-cluster solution was concordant with the pattern observed in the 16S dataset and supported the same broad enterotype method, namely the Enterococcus-type (E-type), Bacteroides-type (B-type), and Ruminococcus-type (R-type). Together, these results indicate that the VAE-based approach recovered a robust and cross-platform three-enterotype structure, with metagenomic data independently validating the major community configurations identified from 16S profiles.

### 3.3. Association of inferred enterotype-like community types with Parkinson’s disease status

Across the three inferred enterotype-like states, the proportion of PD samples was similar: 58.3% in Enterotype 1 (365/626, *P* = 0.776), 60.0% in Enterotype 2 (250/417, *P* = 0.691), and 58.9% in Enterotype 3 (538/914, *P* = 0.973). Thus, no significant difference in PD proportion was observed among the three enterotype-like states.

We next tested whether inferred enterotype-like community types were associated with Parkinson’s disease (PD) status after accounting for study-level heterogeneity. Using the merged dataset of 1,957 samples from six cohorts, we fitted a generalized linear mixed-effects model (GLMM) with cohort included as a random intercept and Enterotype 3 specified as the reference category. The overall contribution of enterotype to PD status was not significant in the primary model (likelihood ratio test: χ² = 2.56, df = 2, *P* = 0.278; Table S2), indicating that inferred enterotype-like structure did not significantly improve discrimination between PD and healthy controls. The main effect estimates from the primary and sensitivity models are summarized in Table 2, with the full model coefficients provided in Table S1.

**Table 2.** Association of inferred enterotype-like community types with Parkinson’s disease status in the primary and sensitivity models *Note.* Primary GLMM, generalized linear mixed-effects model with cohort as a random intercept. Sensitivity GLM, fixed-effects logistic regression adjusted for country. Enterotype 3 was specified as the reference category. Odds ratios (ORs) and 95% confidence intervals (CIs) were obtained by exponentiating model coefficients. The overall contribution of enterotype was not significant in either the primary model (likelihood ratio test: χ² = 2.562, df = 2, P = 0.278) or the sensitivity model (analysis of deviance: χ² = 1.279, df = 2, P = 0.528).

| Model | Comparison | $\beta$ | SE | OR | 95% CI | $P$ value |
| --- | --- | --- | --- | --- | --- | --- |
| Primary GLMM | Enterotype 1 vs 3 | -0.213 | 0.144 | 0.808 | 0.61–1.07 | 0.138 |
| Primary GLMM | Enterotype 2 vs 3 | -0.183 | 0.149 | 0.833 | 0.62–1.12 | 0.220 |
| Sensitivity GLM | Enterotype 1 vs 3 | -0.104 | 0.133 | 0.902 | 0.69–1.17 | 0.437 |
| Sensitivity GLM | Enterotype 2 vs 3 | -0.160 | 0.146 | 0.852 | 0.64–1.14 | 0.275 |

*Note.* Primary GLMM, generalized linear mixed-effects model with cohort as a random intercept. Sensitivity GLM, fixed-effects logistic regression adjusted for country. Enterotype 3 was specified as the reference category. Odds ratios (ORs) and 95% confidence intervals (CIs) were obtained by exponentiating model coefficients. The overall contribution of enterotype was not significant in either the primary model (likelihood ratio test: $\chi^2 = 2.562$ , $df = 2$ , $P = 0.278$ ) or the sensitivity model (analysis of deviance: $\chi^2 = 1.279$ , $df = 2$ , $P = 0.528$ ).

At the level of individual enterotype contrasts, neither Enterotype 1 nor Enterotype 2 showed a significant association with PD in the primary GLMM (Table 2). Relative to Enterotype 3, Enterotype 1 had an estimated coefficient of −0.213 (SE = 0.144, *P* = 0.138), corresponding to an odds ratio (OR) of approximately 0.81, while Enterotype 2 had an estimated coefficient of −0.183 (SE = 0.149, *P* = 0.220), corresponding to an OR of approximately 0.83. Although both estimates were directionally below 1, their confidence intervals overlapped the null, providing no evidence for an enrichment or depletion of specific inferred enterotypes in PD after controlling for cohort effects. The random-intercept variance for cohort was 0.0816 (SD = 0.286), supporting the presence of modest residual between-cohort heterogeneity (Table S1).

We then performed a sensitivity analysis using a generalized linear model (GLM) in which country was included as a fixed effect. This analysis yielded the same overall conclusion: enterotype remained non-significant at the overall level (analysis of deviance: χ² = 1.28, df = 2, *P* = 0.528; Table S2). Relative to Enterotype 3, Enterotype 1 showed an estimated coefficient of −0.104 (*P* = 0.437; OR = 0.90, 95% CI: 0.69–1.17), and Enterotype 2 showed an estimated coefficient of - 0.160 (*P* = 0.275; OR = 0.85, 95% CI: 0.64–1.14) (Table 2 and Table S1). In the sensitivity GLM, country effects were significant for Japan (β = 0.417, *P* = 0.013) and the USA (β = 0.532, *P* < 0.001), whereas Germany and Korea were not significant (Table S1). In this model, which simultaneously adjusted for both country and enterotype, some levels of the country variable remained significantly associated with PD status, whereas the overall effect of enterotype was still non-significant.

Together, these results showed that enterotype assignment was not significantly associated with PD status in either the primary mixed-effects model or the country-adjusted sensitivity model.

### 3.4. Conventional machine-learning benchmarks for PD diagnosis

We next evaluated six conventional machine-learning classifiers as reference models for PD diagnosis. Receiver operating characteristic analysis based on aggregated out-of-fold diagnosiss showed that Random Forest yielded the highest discriminative performance (AUC = 0.775), with Gradient Boosting ranking second (AUC = 0.747). The remaining models performed substantially less well, including Logistic Regression (AUC = 0.654), kNN (AUC = 0.652), Linear SVM (AUC = 0.647), and Neural Network (AUC = 0.625) in Figure 5. The same overall ranking was also observed in the mean ROC analysis, supporting the robustness of this benchmark comparison. Table S3 reports mean accuracy, mean AUC, and corresponding 95% confidence intervals estimated across repeated stratified evaluations.

**Figure 5.**
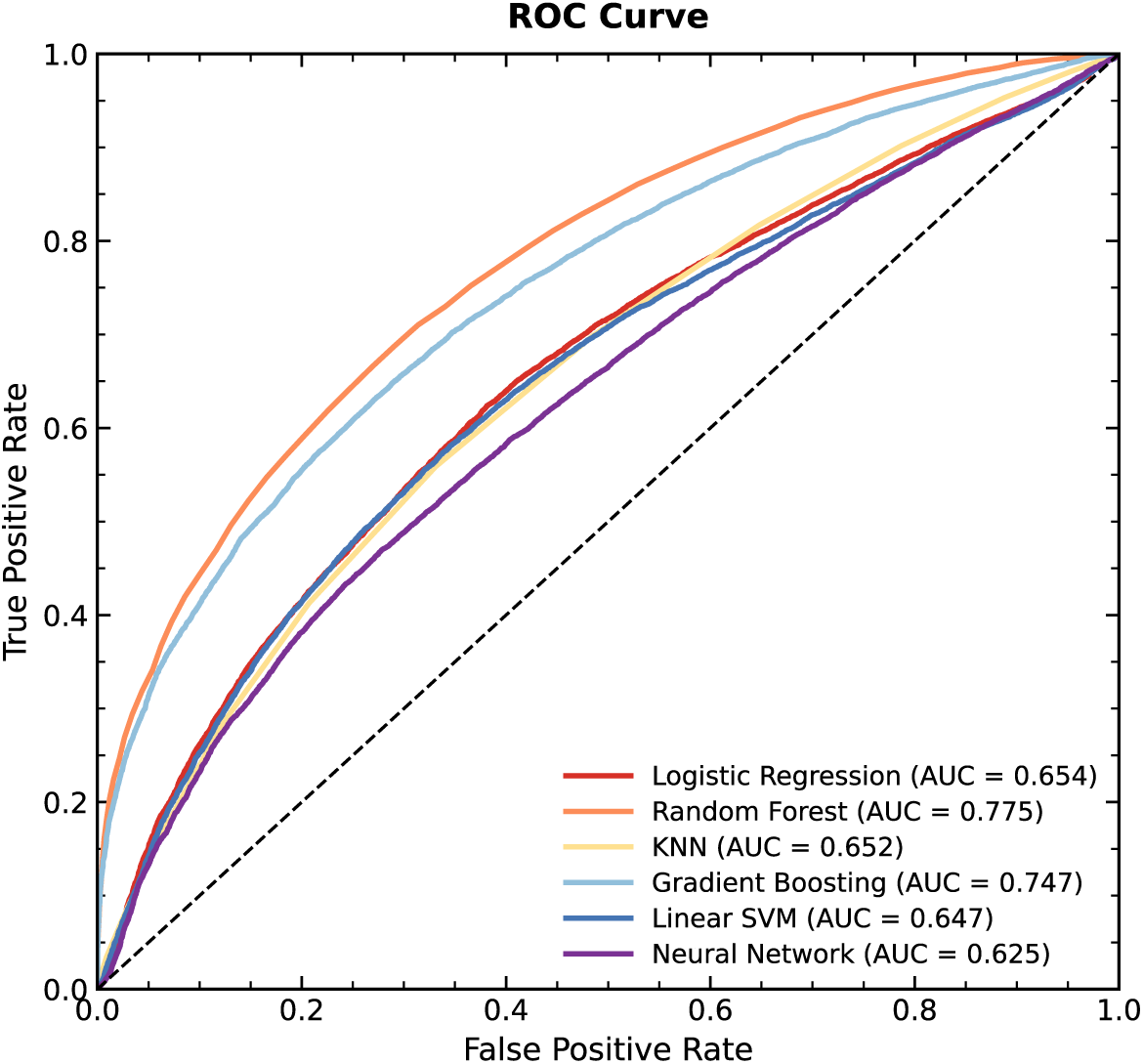
Performance comparison of six baseline machine-learning models for Parkinson’s disease diagnosis. Receiver operating characteristic (ROC) curves were generated from aggregated out-of-fold diagnosis across repeated stratified evaluations. Among the six baseline classifiers, Random Forest achieved the highest discriminative performance, followed by Gradient Boosting, whereas Logistic Regression, k-nearest neighbors (kNN), Linear Support Vector Machine (SVM), and Neural Network showed comparatively lower predictive performance. AUC values are shown in the legend for each model.

### 3.5. Fine-tuning of the SupVAE-based diagnosis branch

We next optimized the SupVAE-based diagnosis branch by systematically varying latent dimensionality, regularization strength, classification-loss weighting, selected feature number, and downstream classifiers. Detailed benchmarking results are provided in Table S4 and Fig S3. TableS4 summarizes predictive performance across all tested SupVAE-based model configurations. Hyperparameters included latent dimensionality, KL divergence penalty (β), classification-loss weight (α), number of selected microbiome features, and downstream classifier. For each configuration, mean performance metrics and corresponding 95% confidence intervals were estimated across repeated stratified cross-validation. Models were ranked primarily by mean AUC to identify the optimal parameter setting for subsequent validation analyses.

### 3.6. Cross-cohort validation of SupVAE-based diagnosis models

Under strict leave-one-cohort-out (LOCO) validation, pooled ROC analysis of all held-out predictions yielded an AUC of 0.660 for Raw RF, 0.655 for SupVAE + RF, and 0.657 for SupVAE + Enterotype + RF (Fig. 6). Thus, the pooled ROC curves showed closely similar discrimination across the three models.

**Figure 6.**
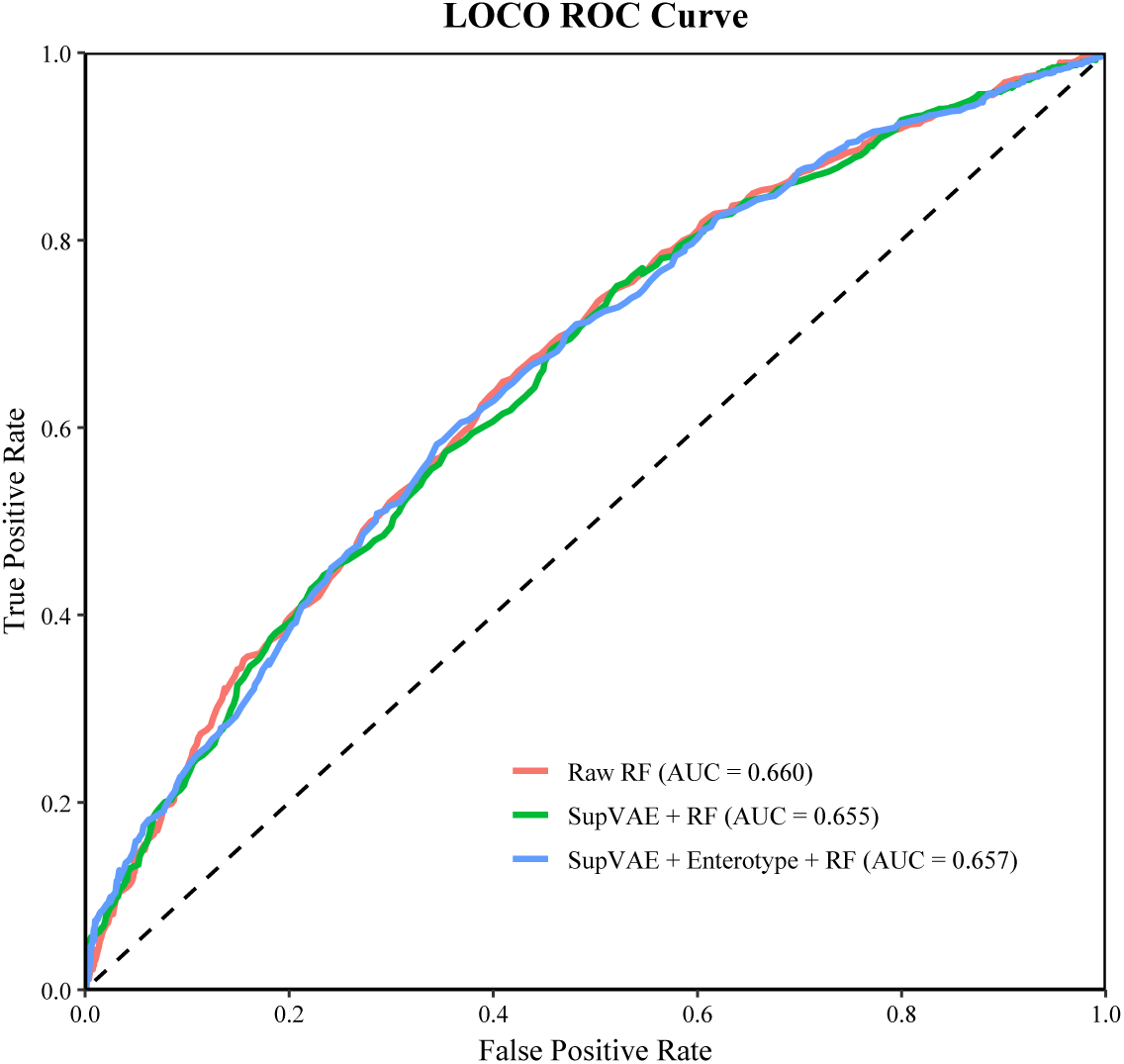
Strict leave-one-cohort-out (LOCO) validation of baseline and SupVAE-based models for Parkinson’s disease diagnosis. Receiver operating characteristic (ROC) curves were generated from pooled held-out predictions across all cohorts under strict leave-one-cohort-out (LOCO) validation. Three models were compared: Random Forest trained directly on filtered microbiome features (Raw RF), a SupVAE-based hybrid model using latent representations plus selected microbiome features (SupVAE + RF), and a SupVAE-based hybrid model further augmented with predicted enterotype labels assigned by nearest-centroid mapping in the training latent space (SupVAE + Enterotype + RF). AUC values shown in the legend represent pooled discrimination across all held-out samples rather than the arithmetic mean of cohort-specific AUCs.

When performance was summarized at the cohort level, the mean AUC was 0.634 (95% CI: 0.524–0.745) for Raw RF, 0.637 (95% CI: 0.553–0.721) for SupVAE + RF, and 0.634 (95% CI: 0.536–0.733) for SupVAE + Enterotype + RF. Mean accuracy values were 0.564, 0.561, and 0.559, respectively. Mean balanced accuracy values were 0.515 for Raw RF, 0.508 for SupVAE + RF, and 0.503 for SupVAE + Enterotype + RF. Mean sensitivity values were 0.994, 0.995, and 0.996, whereas mean specificity values were 0.037, 0.021, and 0.010, respectively (Table S5). Table S5 summarizes model performance across held-out cohorts under strict LOCO validation. For each model, mean AUC, accuracy, balanced accuracy, sensitivity, specificity, and F1 score are reported together with 95% confidence intervals. These values represent cohort-level averages.

Performance varied across individual held-out cohorts (Table S6). The highest cohort-specific AUC was observed for Raw RF in PRJEB30615 (AUC = 0.731), followed by SupVAE + Enterotype + RF in the same cohort (AUC = 0.719) and SupVAE + RF in the same cohort (AUC = 0.713). In PRJDB8639, AUC values were 0.674 for Raw RF, 0.671 for SupVAE + RF, and 0.677 for SupVAE + Enterotype + RF. In PRJEB27564, the corresponding AUC values were 0.609, 0.613, and 0.623. In PRJNA381395, all three models showed lower discrimination, with AUC values of 0.435 for Raw RF, 0.488 for SupVAE + RF, and 0.454 for SupVAE + Enterotype + RF. In PRJNA601994, AUC values were 0.675, 0.666, and 0.666, respectively. In PRJNA742875, AUC values were 0.682, 0.671, and 0.667, respectively. Table S6 reports cohort-specific results for Raw RF, SupVAE + RF, and SupVAE + Enterotype + RF under strict LOCO validation. For each held-out cohort, the table provides sample size, class composition, selected classification threshold, AUC, accuracy, balanced accuracy, sensitivity, specificity, and F1 score.

For SupVAE + Enterotype + RF, held-out enterotype labels were reassigned by nearest-centroid mapping in the latent space constructed from the training cohorts (Table S7). Across all held-out samples, the agreement between reassigned and original enterotype labels was 54.5% (1066/1957).

Cohort-specific agreement rates were 61.4% for PRJDB8639, 66.5% for PRJEB27564, 5.1% for PRJEB30615, 92.1% for PRJNA381395, 60.0% for PRJNA601994, and 54.1% for PRJNA742875.

For the SupVAE + Enterotype + RF model, enterotype labels for held-out samples were not taken directly from metadata but were reassigned by nearest-centroid mapping in the latent space derived from the training cohorts. Table S7 lists the original and reassigned enterotype labels for each held-out sample and supports evaluation of enterotype transferability across cohorts.

## 4. Discussion

In this study, we developed a VAE-based methodology for deep enterotyping in Parkinson’s disease (PD) gut microbiome data and evaluated its performance in comparison with conventional enterotype pipelines and standard machine-learning classifiers. Several conclusions emerge from the results. First, the merged PD microbiome dataset contained community structure that was detectable but not well resolved by classical enterotyping approaches. PAM recovered a nominal three-cluster solution, yet the low silhouette score and diffuse ordination boundaries indicated weak separation. DMM captured finer heterogeneity, but the 12 inferred components remained strongly overlapping and were better interpreted as a continuous compositional landscape than as discrete microbial states. In contrast, the VAE-derived latent space yielded a clearer three-cluster configuration, supported by improved cluster separation in 16S data and by concordant recovery of the same broad three-enterotype method in the independent metagenomic dataset. Together, these findings indicate that nonlinear latent representation learning can provide a more coherent and biologically interpretable basis for enterotyping than conventional distance-based or probabilistic mixture models when applied to sparse, heterogeneous microbiome profiles.

A key strength of the present methodology lies in its ability to handle the statistical properties of microbiome data more effectively than classical methods. Gut microbial abundance tables are sparse, compositional, and often dominated by nonlinear relationships that are difficult to represent in shallow embeddings. The combination of compositional preprocessing, robust scaling, and VAE-based compression appeared to reduce noise from rare features while preserving higher-order structure relevant to community organization. This is likely why the VAE latent space produced a more compact three-enterotype solution, whereas conventional low-dimensional embeddings yielded inconsistent cluster numbers and uneven separation. Importantly, the VAE methodology did not simply generate mathematically cleaner partitions; it also recovered enterotype assignments that were biologically intelligible, corresponding to an Enterococcus-type, a Bacteroides-type, and a Ruminococcus-type configuration.

The biological interpretation of these three community states is broadly consistent with prior ecological understanding of the gut microbiome. The Bacteroides-type resembled a bile-tolerant, protein- and fat-associated configuration, whereas the Ruminococcus-type was more consistent with fiber degradation and short-chain fatty acid production, although some samples within this group also suggested a more inflammatory mucin-degrading substate. The Enterococcus-type appeared to represent a lower-diversity, dysbiosis-like configuration enriched in opportunistic taxa. Although these labels should not be overinterpreted as sharply bounded ecological entities, their recurrence across 16S and metagenomic data suggests that the VAE methodology captured major axes of gut community organization that are robust to sequencing modality. This cross-platform concordance is particularly important, because it argues that the inferred structure is not merely an artefact of one profiling strategy.

In the present study, enterotype assignment was not significantly associated with Parkinson’s disease (PD) status. This conclusion was supported at multiple levels. First, the proportion of PD samples was similar across the three inferred enterotype-like states, indicating no obvious enrichment of PD within any single coarse community type. Second, in the primary generalized linear mixed-effects model (GLMM), which treated cohort as a random intercept to account for study-level heterogeneity across the merged six-cohort dataset, the overall effect of enterotype on PD status was not significant. Third, the same conclusion was obtained in a sensitivity generalized linear model (GLM). Together, these analyses indicate that, in this dataset, enterotype assignment was not a significant differentiating factor for PD status and that the lack of association was not dependent on a single modelling strategy.

This finding is not inconsistent with previous literature. Following the original introduction of the enterotype concept, Arumugam et al. (2014) later clarified that enterotypes should not be regarded as fixed, strictly discrete disease labels, but rather as a pragmatic framework for simplifying the complexity of the gut microbiome and stratifying samples into broad community configurations. In the sensitivity GLM, country effects were significant for Japan (*β* = 0.417, P < 0.05) and the USA (*β* = 0.532, P < 0.001), whereas Germany and Korea were not significant (Table S1). This pattern is consistent with the meta-analysis by Romano et al. (2021), which showed that although compositional differences between PD and control microbiomes could be detected, the overall effect size was modest and substantial between-study heterogeneity persisted, arguing against a stable enterotype-based case-control signal in PD. In addition, Vascellari et al. (2021) reported associations between enterotype-like microbiota/metabolome configurations and clinical phenotypes within PD, suggesting that these community states may reflect intradisease heterogeneity rather than disease presence per se. Similarly, Devolder et al. (2023) showed in multiple sclerosis that the inflammation-associated Bact2 enterotype was independently associated with long-term disability worsening, indicating that microbiome-based stratification may, in some settings, be more informative for prognosis or clinical subtyping than for binary diagnosis.

Taken together, our findings support the interpretation that, in PD, enterotype-like states primarily capture broad ecological background and inter-individual heterogeneity rather than serving as disease-specific markers. It should also be noted that our dataset lacked sufficiently detailed PD subtype information, including tremor-dominant and non-tremor-dominant classifications, and we were therefore unable to test whether enterotype assignment was associated with clinical heterogeneity within PD.

Among the conventional classifiers, Random Forest showed the strongest baseline discrimination, indicating that nonlinear models are better suited than linear classifiers to the complexity, sparsity, and compositional structure of PD gut microbiome data. Against this benchmark, the supervised VAE-based prediction branch achieved cross-cohort performance that was broadly comparable to the raw-feature Random Forest model. Although this meant that the VAE framework did not deliver a large gain in predictive accuracy alone, its methodological value should not be judged solely on that basis. Rather, the key advantage of the VAE approach lies in its ability to provide a shared latent representation that links unsupervised enterotype inference with supervised case–control prediction within the same analytical framework.

This shared representation is important for several reasons. First, it allows clustering, out-of-sample enterotype transfer, and disease classification to be interpreted with reference to the same underlying structure, thereby improving coherence across analytical tasks that are usually treated separately. Second, it offers a lower-dimensional and more organized representation of highly sparse, noisy, and compositional microbiome data, which is particularly useful for cross-cohort comparison and for reducing the instability that often arises when raw features are analysed directly. Third, because the same latent space can be carried forward into downstream analyses, the framework facilitates biologically informed stratification and interpretation of broad community states in a way that conventional raw-feature classifiers do not naturally support. In this sense, the principal contribution of the VAE-based methodology is not simply to maximize classification performance, but to establish a reproducible, transferable, and interpretable latent structure that can support multiple downstream analyses within one coherent model.

Several limitations should be acknowledged. First, this was a retrospective multi-cohort integration study, and substantial heterogeneity remained despite harmonized processing and explicit adjustment for cohort or country. Important clinical covariates such as bowel habits, medication exposure, disease duration, diet, and disease severity were not uniformly available across datasets, and residual confounding is therefore likely. Second, the enterotype method adopted here was based on hard assignment, which is useful for interpretation and downstream modelling but may simplify a microbial landscape that is at least partly continuous. Third, the metagenomic validation analysis supported the same three-enterotype structure, but it was based on a single external shotgun cohort; broader validation across additional metagenomic datasets will be needed to establish generalizability more firmly. Fourth, our analyses were performed primarily at the genus level. This improves harmonization across cohorts and platforms, but it may obscure strain-level, pathway-level, or gene-level signals that are more directly relevant to PD biology. Finally, transferability of enterotype labels across cohorts was imperfect in some held-out settings, indicating that even a biologically meaningful latent structure remains sensitive to cross-study distribution shifts.

These limitations also point to the next steps. Future work should test the method in prospectively collected cohorts with richer metadata, evaluate whether enterotype-stratified analyses improve the detection of PD-associated taxa or functions, and extend the model to semi-supervised or covariate-aware architectures that can explicitly partition disease signals from geographic and clinical confounders. It will also be valuable to move beyond taxonomic composition and examine whether metabolomic, functional, or host inflammatory profiles align more strongly with the VAE-defined enterotypes than case–control status alone. Such extensions may clarify whether community types act primarily as ecological backgrounds or as modifiers of disease expression, treatment response, or symptom subphenotypes.

Overall, the present study supports a practical reformulation of enterotyping in PD microbiome research. Rather than asking whether canonical enterotypes exist in a strict and universal sense, the more useful question may be whether representation learning can recover stable, interpretable community states that help organize heterogeneous microbiome variation. Our results suggest that a VAE-based latent method moves the field closer to that goal.

## 5. Conclusion

First, we developed a VAE-based deep enterotyping methodology for PD gut microbiome data and showed that it recovered a clearer and more reproducible three-enterotype structure than conventional PAM, DMM, and standard low-dimensional embedding approaches. The resulting Enterococcus-, Bacteroides-, and Ruminococcus-type community states were biologically interpretable and were independently recapitulated in metagenomic data, supporting the robustness of the inferred broad community structure across profiling platforms.

Second, enterotype was not a significant differentiating factor for PD status in this heterogeneous multi-cohort dataset. This conclusion was supported at multiple levels: the proportion of PD samples was similar across the three inferred enterotype-like states, with no obvious enrichment of PD in any single coarse community type; the primary generalized linear mixed-effects model, which accounted for cohort-level heterogeneity by including cohort as a random intercept, showed no significant overall effect of enterotype on PD status; and the same result was reproduced in the sensitivity generalized linear model.

Finally, the prediction branch showed that VAE-derived representations could support PD case–control classification while also providing a shared latent representation for clustering, enterotype transfer, and downstream interpretation. This indicates that the value of the supervised branch lies not only in predictive modelling itself, but also in linking disease classification with the same latent structure used for enterotype inference.

In conclusion, this study is the first to apply a VAE-based deep learning framework to enterotype inference in Parkinson’s disease gut microbiome data. Our results show that this approach provides a robust strategy for PD enterotyping and highlights the value of deep representation learning for resolving broad community structure in complex compositional datasets. More broadly, the findings suggest that the principal utility of enterotypes in PD may lie in reproducible community stratification and cross-cohort organization, rather than in direct diagnostic discrimination alone.

## Ethics statement

This study was based exclusively on previously published and publicly available de-identified sequencing data retrieved from the National Center for Biotechnology Information (NCBI). No new human participants were recruited, and no new samples or identifiable personal data were collected for this study. Ethical approval and informed consent were obtained by the original studies, as described in their respective publications. Therefore, no additional institutional ethics approval was required for this secondary analysis of publicly available data.

## Data availability

All raw sequencing data analyzed in this study are publicly available from the National Center for Biotechnology Information (NCBI) under the following BioProject accessions: PRJDB8639, PRJEB27564, PRJEB30615, PRJNA381395, PRJNA601994, PRJNA742875, and PRJNA834801.

Dataset-level metadata were curated from the corresponding original publications. The processed genus-level abundance tables generated in this study are available from the corresponding author upon reasonable request.

## Data Availability

Data availability
All raw sequencing data analyzed in this study are publicly available from the National Center for Biotechnology Information (NCBI) under the following BioProject accessions: PRJDB8639, PRJEB27564, PRJEB30615, PRJNA381395, PRJNA601994, PRJNA742875, and PRJNA834801. Dataset-level metadata were curated from the corresponding original publications. The processed genus-level abundance tables generated in this study are available from the corresponding author upon reasonable request.

